# Validation of the Incremental Prognostic Value of Deceased Donation Pre-implantation Kidney Transplant Biopsies through Comprehensive Lesion Quantification with Deep Learning

**DOI:** 10.64898/2026.09.08.26362425

**Authors:** Ziyang Li, Giorgio Buzzanca, Danny van der Helm, Soufian Meziyerh, Ian P.J. Alwayn, Dorottya K. de Vries, Hans J. Baelde, Aiko P.J. de Vries, Jesper Kers

## Abstract

**Background:** Pre-implantation biopsy utility for deceased-donor kidney acceptance is controversial due to processing limitations and interpretive variability. We evaluated whether systematic histological assessment, including expert evaluation and quantification with the BanffNET automated deep learning system (17 lesions), predicts post-transplant failure under optimal laboratory conditions.

**Methods:** Biopsies from deceased donor kidneys (N=733) underwent optimized laboratory processing, whole slide imaging, expert Banff 2024 pathologic assessment, and BanffNET lesion quantification. We assessed their incremental predictive value beyond clinical characteristics for early graft dysfunction, longitudinal eGFR/UPCR trajectories, need for indication biopsies, and death-censored graft failure.

**Results:** Histopathology improved clinical model discrimination by 1-4% and increased explained variance for longitudinal trajectories by 1-2%. Continuous bivariate surface analysis, however, revealed that while histology adds marginal overall predictive accuracy, mapping continuous composite and individual BanffNET scores against the Kidney Donor Risk Index (KDRI) uncovers distinct topological risk gradients for early graft dysfunction and long-term graft failure.

**Conclusion:** While pre-implantation biopsies add minimal overall prognostic value, continuous bivariate mapping of KDRI and BanffNET scores uncovers distinct synergistic risk gradients for graft failure. These continuous clinical-histological surfaces could optimize organ salvage by precisely identifying viable high-KDRI kidneys lacking critical compounded damage and vice versa.

**Lay Summary:** Kidney transplantation is the optimal treatment for end-stage kidney disease, but donor shortages lead to long waiting times. Some donated kidneys are declined because biopsies taken during allocation show unfavorable findings. We assessed whether these biopsies improve prediction of post-transplant outcomes beyond routinely available donor and recipient characteristics. Using optimized tissue processing, expert kidney pathologist assessment, and BanffNET automated deep-learning analysis of digital whole-slide images, we found that adding biopsy findings resulted in only small improvements in outcome prediction. Thus, even under optimized conditions, pre-implantation biopsies provide limited additional information for assessing donor kidney suitability and may contribute to unnecessary organ discard. Automated digital analysis with BanffNET may help reduce variation between pathologists and improve histological assessment and risk stratification, particularly for kidneys from higher-risk donors.

## Introduction

Kidney transplantation is the optimal treatment option for patients with end-stage kidney disease (ESKD)^1^. However, a persistent imbalance between organ availability and the growing waiting list has existed for decades^2^. To address this shortage, expanded criteria donors (ECD) have increasingly been utilized^3^. Given the underlying risk of ECD kidneys, careful evaluation of organ quality is essential to prevent unnecessary discard while minimizing the risk of primary nonfunction^4^. In addition to the routinely used Kidney Donor Risk Index (KDRI)^5^, some transplant centers rely on donor kidney biopsies to evaluate organ quality^6^.

Currently, three main types of pretransplant biopsies are being performed: (1) procurement biopsies, which are used to assess organ quality and guide acceptance or rejection of allocation decisions^7^; (2) pre-implantation biopsies, which are used to evaluate chronic damage and donor-derived pathology and serve as baseline references for post-transplant indication biopsies^8^; and (3) post-reperfusion biopsies, typically obtained approximately 30 minutes after reperfusion, which are used to assess early acute injury and pre-existing chronic damage^9,10^. Specifically the use of **procurement kidney biopsies** for the purpose of allocation has sparked considerable debate^11^, and several studies suggested that biopsy findings may have contribute to unnecessary organ discard, at least in the United States^12–15^. Corroborating this, the Banff Time-Zero Biopsy Working Group recommended that procurement biopsies should not be performed routinely, but rather when donor clinical characteristics would unlikely lead to a kidney transplantation in such a high-risk setting (organ salvage)^8,16^. The procurement biopsy could then rescue the donor kidney for transplantation.

Disagreements regarding the utility and context of use of procurement biopsies mainly arose from two aspects. First, the reproducibility of pathology reports on such biopsies is limited for multiple reasons that are not mutually exclusive^17^. Frozen-section specimens stained with hematoxylin and eosin (H&E) often suffer from freezing artifacts, which may lead to false-positive assessments of chronic damage. Poor structural resolution of frozen material hampers accurate evaluation of glomerulosclerosis, interstitial fibrosis and tubular atrophy, and chronic vascular changes, while restricted staining options (only H&E) result in incomplete pathological interpretation^18,19^. Furthermore, assessments are frequently performed by on-call pathologists without expertise in renal pathology under time constraints, further increasing interobserver variability and reducing reliability compared to daytime assessment by a pathologist with expertise in renal pathology^20^. Collectively, the assessment of procurement biopsies is being performed under severely constrained circumstances.

Second, data on the prognostic value of performing a procurement biopsy remains inconsistent^21^. This is substantially attributable to the suboptimal assessment procedures described above^22,23^, and often histopathological findings are being interpreted in isolation, rather than in addition to the KDRI.

Unlike retrospective registry studies in the United States, which are heavily constrained by confounding by indication (as biopsies are triggered by high-risk donor clinical profiles) and severe selection bias (since kidneys with high chronicity scores are systematically discarded and never transplanted), our cohort represents a non-selection design. Because pre- implantation biopsies in our center are used strictly as a baseline reference and do not influence allocation decisions, we were able to evaluate the prognostic value of histopathological chronicity across its entire pathological spectrum. This study therefore aims to determine the potential incremental prognostic value pre-implantation biopsies could maximally provide beyond baseline clinical characteristics when optimized histopathological conditions are applied to address the limitations mentioned above (standardization and optimization of the laboratory procedures and the pathologist assessment). We evaluate their association with early graft damage, kidney function trajectories and long-term graft survival, thereby offering a comprehensive assessment of their potential clinical outcome association. We specifically assessed whether state-of-the-art automated and quantitative computational pathology with the novel BanffNET system, which by definition alleviates inter- and intra-observer variability, could further improve assessment within this optimized context.

Importantly, we explicitly did not aim to create a prediction model for graft outcome as this should be done on material collected under the currently available suboptimal conditions in which the procurement biopsies are being performed (i.e. H&E staining on frozen material as per the Banff Time-Zero Biopsy Working Group) and validated as such in external cohorts.

For this reason, we consider the current study as a validation of the performance of histological assessment under optimized laboratory conditions (including quantification with deep learning), indicative of the maximum expected performance under suboptimal conditions with a state-of-the-art deep learning system.

## Methods

### Population and data collection

All kidney transplantations performed at Leiden University Medical Center (LUMC) between January 1, 2011, and June 30, 2025, were eligible for inclusion. Exclusion criteria were as follows: 1) combined kidney–pancreas and kidney–liver transplantations; 2) transplants from living donors (Figure 1). Clinical data were collected prospectively during routine follow-up and stored in electronic medical records linked to LUMC Biobank for Kidney Transplantation (TRAX). A dedicated data manager is in charge of extracting the relevant data for this study.

**Figure 1.**
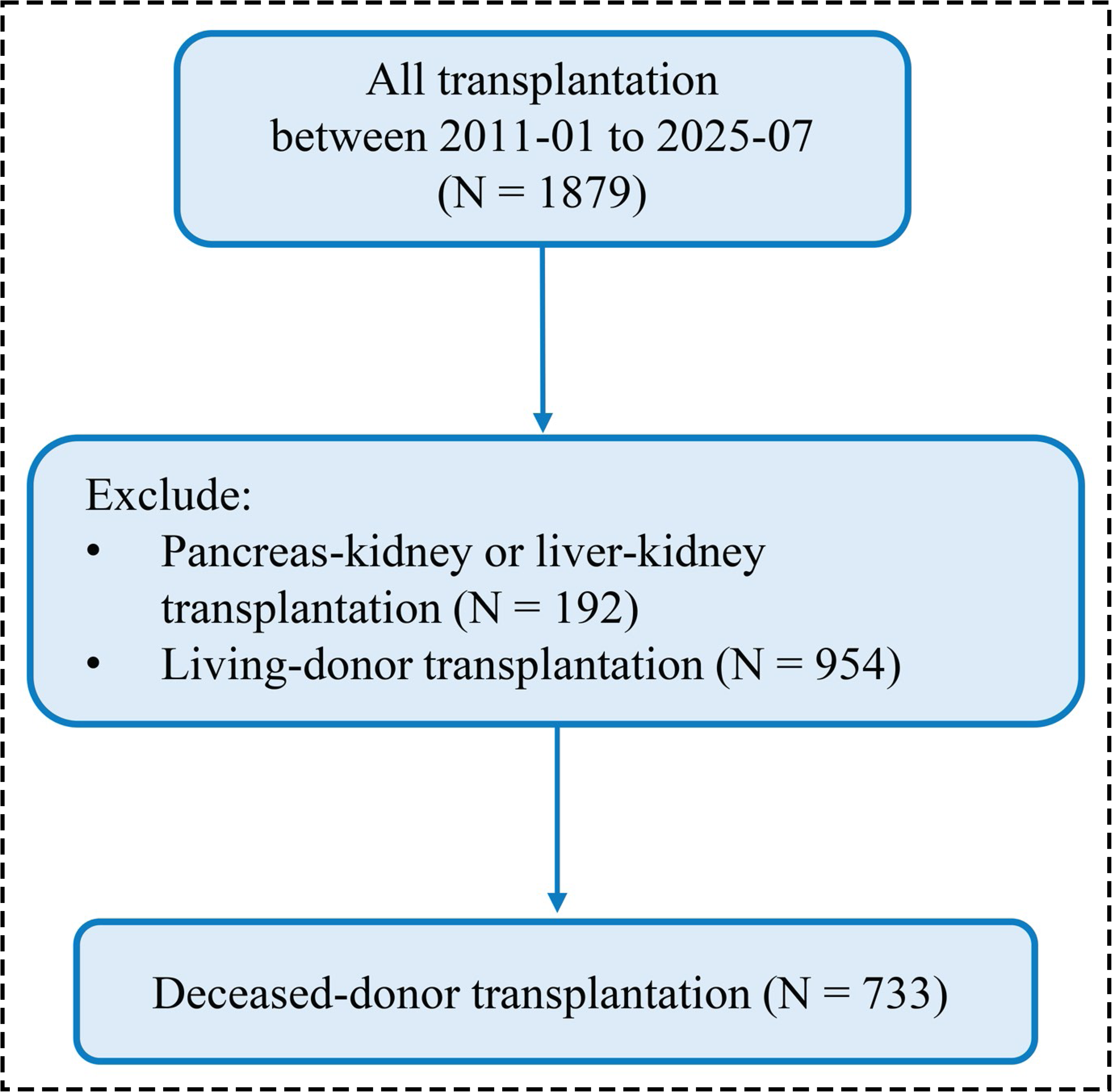
Study workflow. Overview of the study design from patient recruitment to data collection.

This study falls under the approval of the Non-WMO Committee of Leiden University Medical Center, Department of Internal Medicine (ID 138026).

### Optimized histopathological conditions

In the Netherlands, pre-implantation biopsies are primarily used as baseline references rather than a tool for procurement decision making. Wedge biopsies are processed as formalin-fixed, paraffin-embedded (FFPE) sections, enabling highest-quality morphological assessment. In our center, histopathological evaluation is based on three stains (H&E, periodic acid-Schiff (PAS), and Jones methenamine silver (JMS)) and is evaluated by experienced renal pathologists. For this study, all pre-implantation biopsies were retrospectively revised by a single renal pathologist (JK) in a standardized fashion based on the digitized whole slide biopsy images of all three stains according to the latest iteration of the Banff Classification^24^.

### Automated Banff lesion quantification with the BanffNET deep learning algorithm

Within our group we developed an automated, quantitative computational pathology system, called BanffNET^25^. BanffNET has been trained and extensively validated on >20,000 whole-slide images (WSIs) for the following Banff lesion scores: g (extent of glomerulitis), cg (severity of transplant glomerulopathy), mm (severity of mesangial matrix expansion), i (extent of interstitial inflammation), t (severity of tubulitis), ti (extent of total inflammation), ifta (extent of interstitial fibrosis and tubular atrophy), iifta (severity of interstitial inflammation in areas of IFTA), tifta (severity of tubulitis in areas of IFTA), ptc (severity of peritubular capillaritis), v (severity of intimal arteritis), cv (severity of vascular fibrous intimal thickening), ah (severity of arteriolar hyalinosis), tma (presence of thrombotic microangiopathy), ati (presence of acute tubular injury), gs (extent of global glomerulosclerosis and fsgs (extent of focal and segmental glomerulosclerosis). The multimodal BanffNET system produced quantitative lesion scores ranging from 0 to 1 on a continuous scale by integrating quantifications from the three stainings via late fusion. Single stain lesion quantifications are also available, but were not the focus of the current study investigating the added value under the most optimal laboratory circumstances.

### Outcome measures

The incremental value of the pre-implantation biopsy was evaluated across four outcomes. The first outcome was early graft (dys)function, classified as: immediate graft function (IGF), defined as immediate functioning of the transplanted kidney without the need for dialysis during the first 7 days post-transplant; delayed graft function (DGF), defined as the requirement for at least one dialysis session within the first 7 days after transplantation; primary non-function (PNF), defined as irreversible graft failure with no recovery of function, resulting in permanent dialysis dependence, the need for re-transplantation, or graft nephrectomy^26^. The second outcome was the trajectory of kidney function up over 1825 days (5 years), which included longitudinal trajectories of estimated glomerular filtration rate (eGFR), and urine protein-to-creatinine ratio (UPCR). The third outcome was time to the first indication biopsy, representing a clinically relevant post-transplant event independent of the underlying histopathologic diagnosis. This was defined as the time from transplantation to the first kidney allograft biopsy performed for suspected graft injury (indication biopsy), excluding biopsies performed in the setting of PNF and DGF. The fourth outcome was death-censored graft failure (DCGF), defined as return to long-term dialysis or need for kidney re-transplantation at the end of the study (June 30, 2025). In DCGF analysis, recipient deaths with a functioning graft were treated as censored events and not counted as graft failure^26^.

### Kidney Donor Risk Index calculation

The Kidney Donor Risk Index (KDRI) was calculated according to the Organ Procurement and Transplantation Network (OPTN)/United Network for Organ Sharing (UNOS) model using the R package kidney.epi 1.4.0, which applies the published donor-specific regression coefficients from the original KDRI derivation study^5^. Donor variables included age, sex, height, weight, ethnicity, history of hypertension, history of diabetes, cause of death, terminal serum creatinine, hepatitis C virus (HCV) status, and donation after circulatory death (DCD) status.

### Covariate sets

To evaluate the incremental contribution of pre-implantation biopsy-derived variables beyond baseline clinical characteristics, analyses were conducted using predefined covariate sets. Set 1 comprised baseline clinical characteristics, including the KDRI and recipient characteristics: age, sex, height, weight, history of diabetes and hypertension, prior kidney transplantation, HLA mismatch level, and the presence of donor-specific antibodies (DSA) prior to transplantation. We then developed additive models to incorporate histopathological features captured from the pre-implantation biopsy. Set 2 extended Set 1 by incorporating the pathologist-assessed Banff lesion scores and the percentage of glomerulosclerosis. Set 3 extended Set 1 by incorporating all 17 quantitative BanffNET scores (g, cg, mm, i, t, ti, ifta, iifta, tifta, ptc, v, cv, ah, tma, ati, fsgs, and gs). An overview of all included variables can be found in Supplementary Table 1.

Given multicollinearity among several histopathological features in Sets 2 and 3, principal component analysis (PCA) was applied to reduce dimensionality/complexity and summarize correlated predictors. The variance explained by each principal component derived from pathologist-assessed scores and BanffNET-assessed scores is presented in Supplementary file 1. The first seven principal components (PC), explaining 75% of the total variance, were retained for subsequent analyses. PCA analysis reduces *model* complexity, but it has no effect on *lesion scoring* complexity.

Set 4 further extended Set 1 by incorporating principal components derived from the pathologist-assessed scores and Banff lesion scores, whereas Set 5 extended Set 1 by incorporating principal components derived from BanffNET scores. These covariate sets were incorporated sequentially to assess the additional contribution of histopathological features beyond the baseline clinical profile.

### Continuous BanffNET score across the spectrum of KDRI

To validate BanffNET scores across the spectrum of KDRI scores, we created a continuous BanffNET score, defined as the sum of all individual BanffNET scores. The continuous BanffNET score represents the total burden of abnormality of the kidney transplant biopsy. Tis allows us to investigate if BanffNET associates with short- and long-term outcomes in patients with low and high KDRI scores. Bivariate surface plots for early (Non-IGF) and late graft dysfunction (death-censored graft failure) were created to visualize the added value of BanffNET on top of the KDRI.

### Statistical analyses

Variables were summarized using descriptive statistics. Continuous variables were reported as mean ± SD or median (interquartile range when skewed), and categorical variables as counts and percentages. Correlations among clinical characteristics, pathologist-assessed scores, and BanffNET scores were assessed using Spearman rank correlation tests.

Associations between variables and early graft outcomes were evaluated using logistic regression, with IGF as the reference category. Odds ratios (ORs) and 95% confidence intervals (CIs) were reported. Trajectories of kidney function were analyzed using linear mixed-effects models, incorporating random intercepts to account for within-subject correlation. Random slopes for time were included where appropriate. To account for the relationship between longitudinal kidney function and DCGF, joint models for longitudinal and time-to-event data were constructed. With the joint model we could calculate the additive value of baseline risk (clinical set +/- histology) *conditional* on the posttransplant kidney function trajectories, i.e. answer the question if baseline risk factors (either or not including histology) associate with DCGF beyond their association with graft function trajectories. The joint modeling framework consisted of (1) a linear mixed-effects submodel with natural spline functions of time to model longitudinal eGFR and UPCR trajectories and (2) a Cox proportional hazards submodel for the requirement for an indication biopsy and death-censored graft failure, linked through the subject-specific longitudinal biomarker trajectories. Kaplan–Meier analysis with log-rank testing was additionally performed to evaluate graft survival across risk stratification, while Cox proportional hazards regression was performed to estimate hazard ratios (HRs) and 95% confidence intervals (CIs). Missing data were handled using multiple imputation by chained equations, and pooled estimates were combined according to Rubin’s rules. The multivariable models for Non-IGF and DCGF were used as input for the bivariate surface plots.

Model performance was evaluated and compared between the covariate set 1 (clinical baseline profile) and the extended sets (clinical baseline profile together with additional histopathological features). Discriminative ability was assessed using the area under the receiver operating characteristic curve (AUC). Improvement in explained variance was assessed using marginal R² for mixed-effects models. Predictive performance of the joint models was additionally evaluated using the time-dependent Brier score, with lower values indicating better predictive accuracy. Overall model fit and parsimony were evaluated using the Akaike Information Criterion (AIC), Deviance Information Criterion (DIC), and

Watanabe–Akaike information criterion (WAIC), with lower values indicating better fit. Bayesian model evidence was additionally assessed using the posterior marginal log-likelihood (PML), with higher (i.e., less negative) values indicating that the model provided a better explanation of the observed data. Likelihood ratio tests (LRT) were used for model comparisons in logistic regression and linear mixed-effects models.

All analyses were conducted in R (version 4.2.1; R Foundation for Statistical Computing, Vienna, Austria) using the following R packages: nnet 7.3.19, lme4 1.1.37, JMbayes2 0.6.0, pROC 1.19.0.1, transplantr 0.2.0, mice 3.18.0, ggplot2 4.0.2, tidyverse 2.0.0.

## Results

### Study population characteristics

This study included 733 deceased-donor kidney transplants, with a median follow-up of 1,444 days (4.0 years). The mean KDRI was 1.57, and the mean recipient age was 58 years. A total of 15% of recipients had undergone a prior kidney transplantation, and 6.8% were donor-specific antibody (DSA) positive at the time of transplantation. The mean of HLA mismatches level was 3. Early graft function was classified as IGF in 387 patients, DGF in 327 patients, and PNF in 19 patients. For death-censored graft failure, 104 patients (14%) experienced graft failure before the end of follow-up at a median of 646 days (1.8 years) after transplantation. Pathologist-assessed scores were predominantly 0 or 1, and BanffNET scores were similarly concentrated at low values with a right-skewed distribution. Given the extremely low prevalence of ptc, cg, and v lesions (≤1 patient with a score >0), these variables were omitted from subsequent analyses (Table 1, Supplementary Figure 1).

**Table 1.**
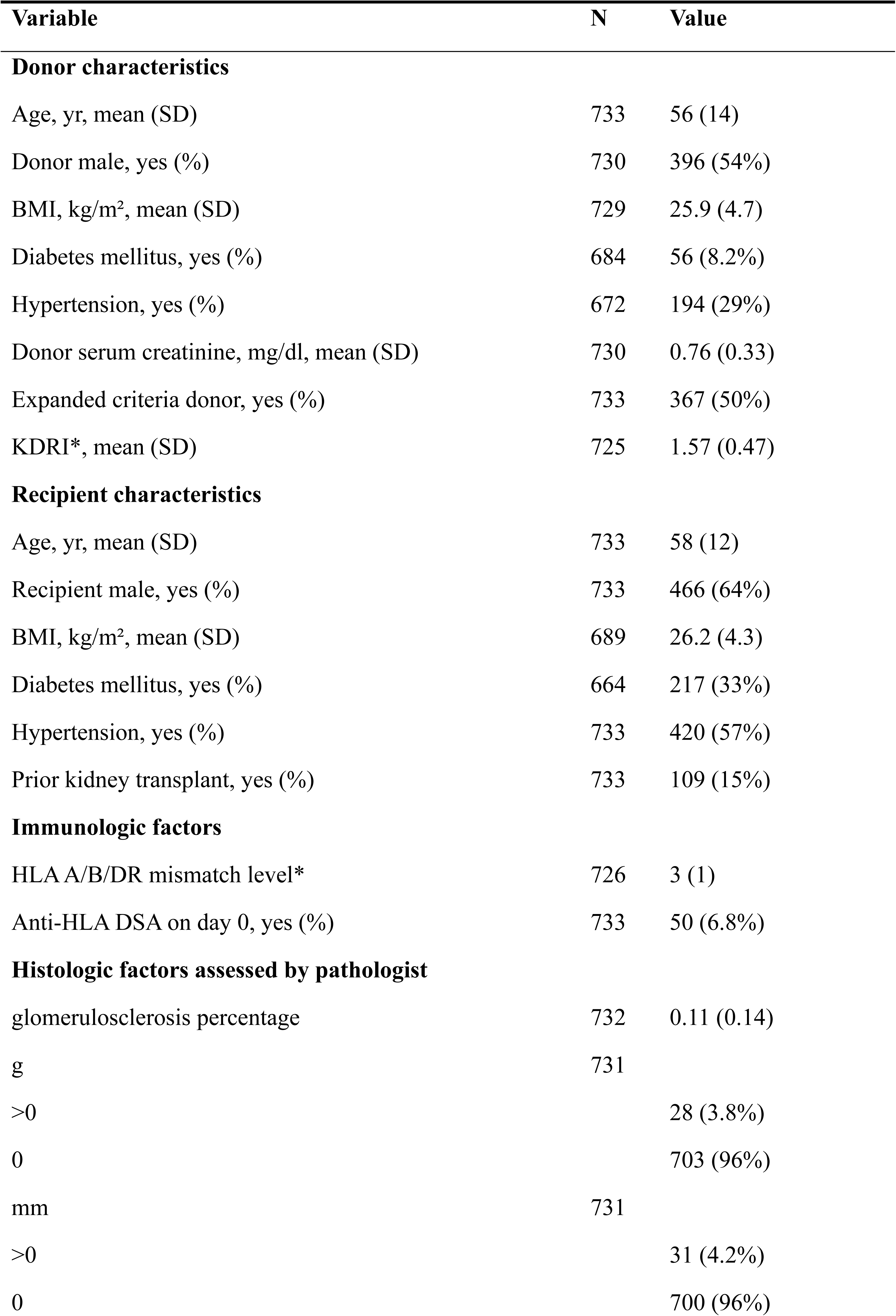
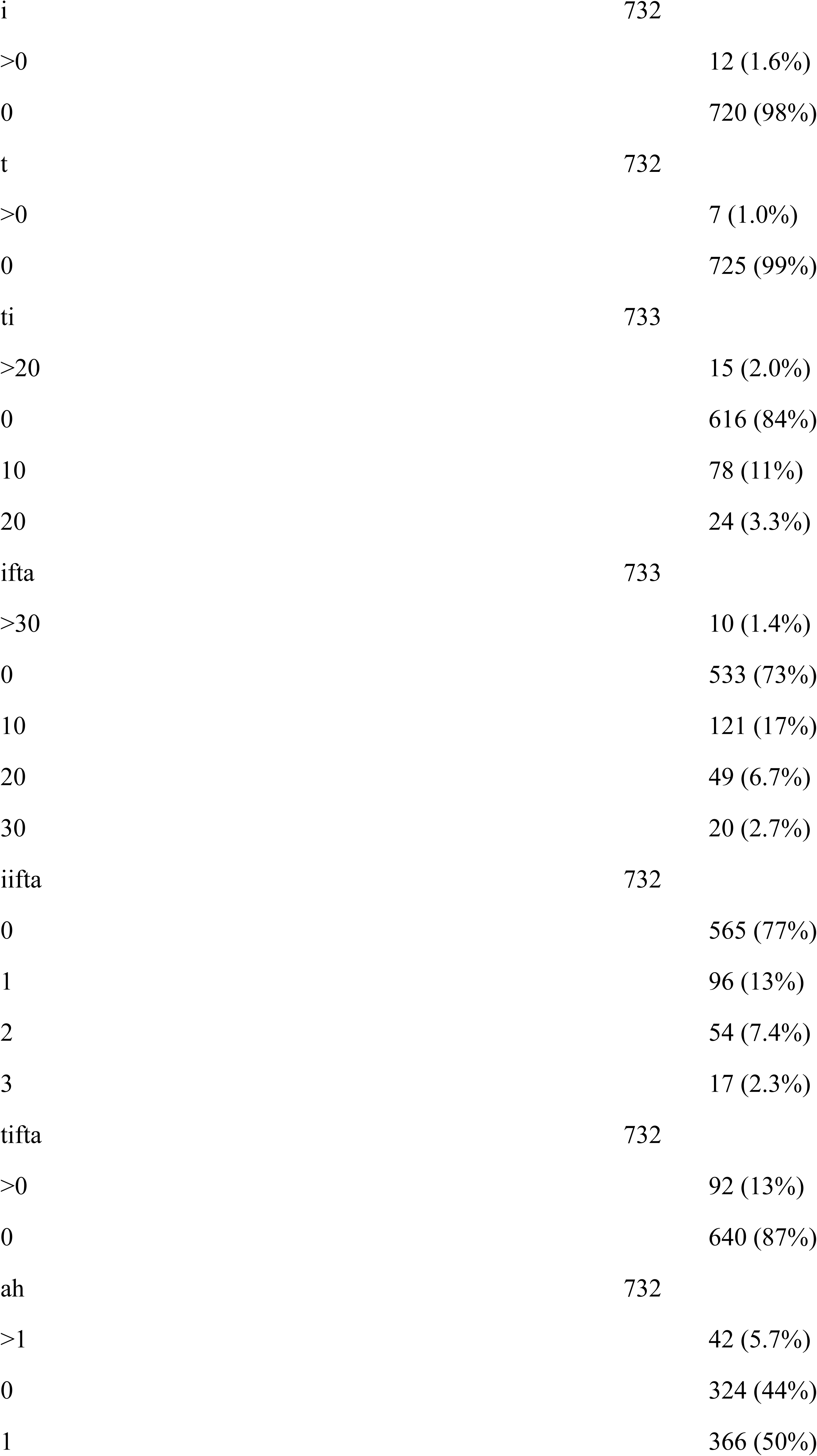
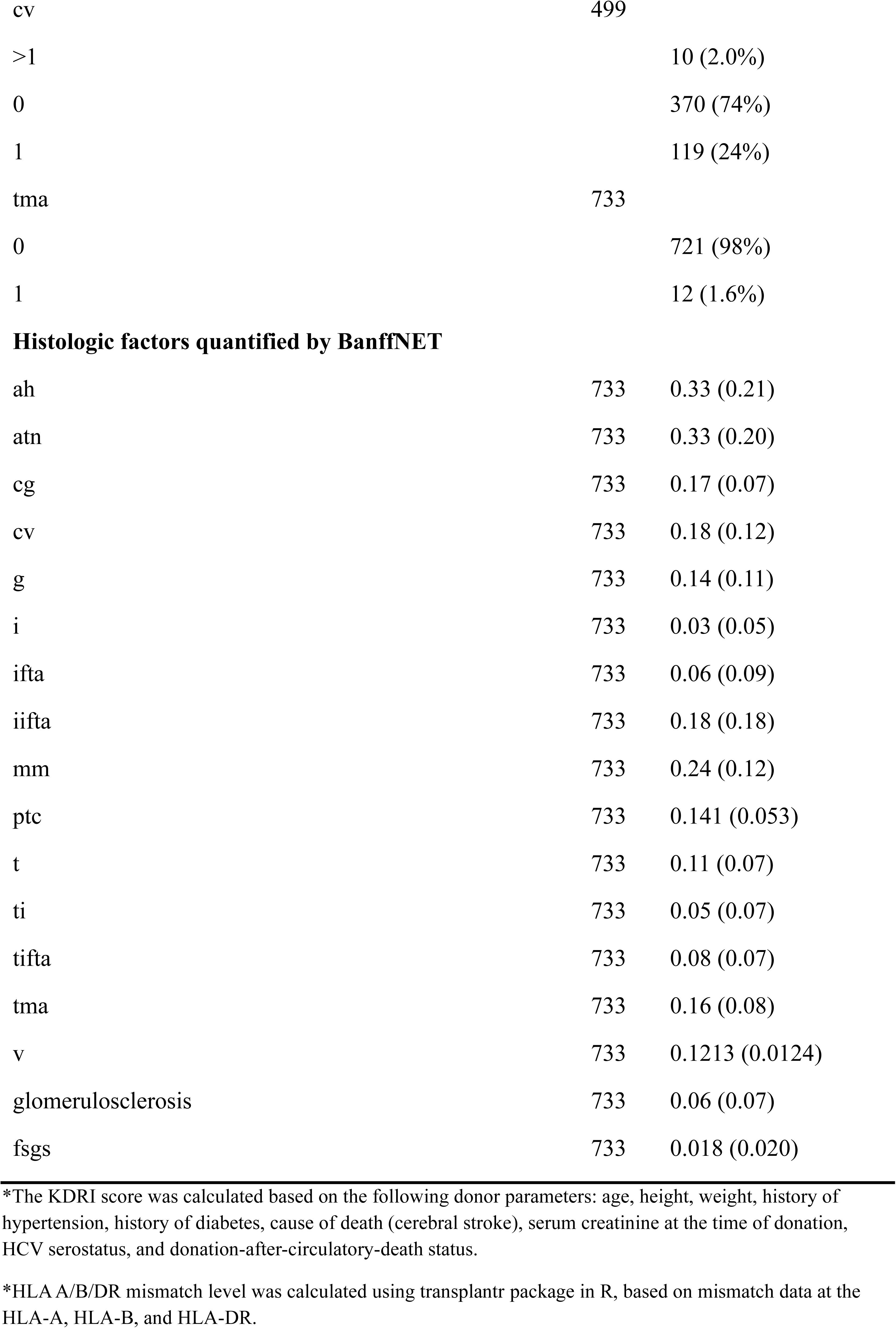
Characteristics of donor, recipient, and biopsy.

Spearman correlations between histopathological features and KDRI are shown in Supplementary Figure 2. Among the pathologist-assessed scores, gs demonstrated the strongest association with KDRI (Spearman ρ = 0.47, p < 0.001 ), whereas the remaining features showed weak or non-significant associations. BanffNET scores demonstrated generally stronger and more consistent correlations, particularly for ifta (ρ = 0.51, p < 0.001 ), ti (ρ = 0.44, p < 0.001 ), and gs (ρ = 0.41, p < 0.001 ), with most associations reaching statistical significance, except for atn. Within-group Spearman correlation matrices of histopathological features were shown in Supplementary Figure 3. Pathologist-assessed scores demonstrated strong correlations primarily among fibrosis-related tubulointerstitial features (e.g., ifta, iifta, tifta; ρ up to 0.89), with generally weaker correlations across other domains. BanffNET scores showed more strong correlation particularly among tubular inflammation and fibrosis features (ti, ifta, iifta), as well as a strong correlation between vascular features (ah and cv).

### Incremental value of histopathological features in predicting early graft (dys)function

Associations of baseline clinical characteristics +/- histopathological features with early graft function (DGF+PNF vs IGF) are presented in Supplementary Figure 4. Compared with the baseline clinical model (Set 1), extending the model with histopathological features generally did not significantly improve model performance. Only the model incorporating the PCs of BanffNET scores showed a statistically significant improvement over the baseline clinical model based on the LRT (p = 0.03). Consistent with this finding, this was also the only extended model associated with improved fit, as reflected by a reduction in AIC (ΔAIC = −1.27), whereas all other extended models increased model complexity without improving fit (positive ΔAICs; Table 2).

**Table 2.** Binary logistic regression analysis for early graft function, model performance comparison.

| Comparison | Extended variables | P value (LRT test) | Δ AIC | Δ Accuracy | Δ AUC |
| --- | --- | --- | --- | --- | --- |
| Set 2 vs Set 1 | Pathologist scores | 0.18 | 7.57 | 0.01 | 0.02 |
| Set 3 vs Set 1 | BanffNET scores | 0.27 | 13.95 | 0.05 | 0.04 |
| Set 4 vs Set 1 | PC of pathologist scores | 0.21 | 4.41 | 0.00 | 0.01 |
| Set 5 vs Set 1 | PC of BanffNET scores | 0.03 | -1.27 | 0.03 | 0.03 |

LRT, likelihood ratio test; AIC, Akaike Information Criterion; AUC, area under the receiver operating characteristic curve; PC, principal component. Δ indicates the change compared with the baseline model (Set 1). Set 1 represents the Baseline model, while Sets 2–5 include additional covariates as specified. A p-value < 0.05 was considered statistically significant.

Model discrimination for early graft dysfunction was evaluated using ROC analysis (Figure 2). Compared with the baseline clinical model, the extended models showed only modest improvements in discrimination, with ΔAUC ranging from 0.01 to 0.04. Similarly, model accuracy improved only marginally, with Δaccuracy ranging from 0.00 to 0.05.

**Figure 2.**
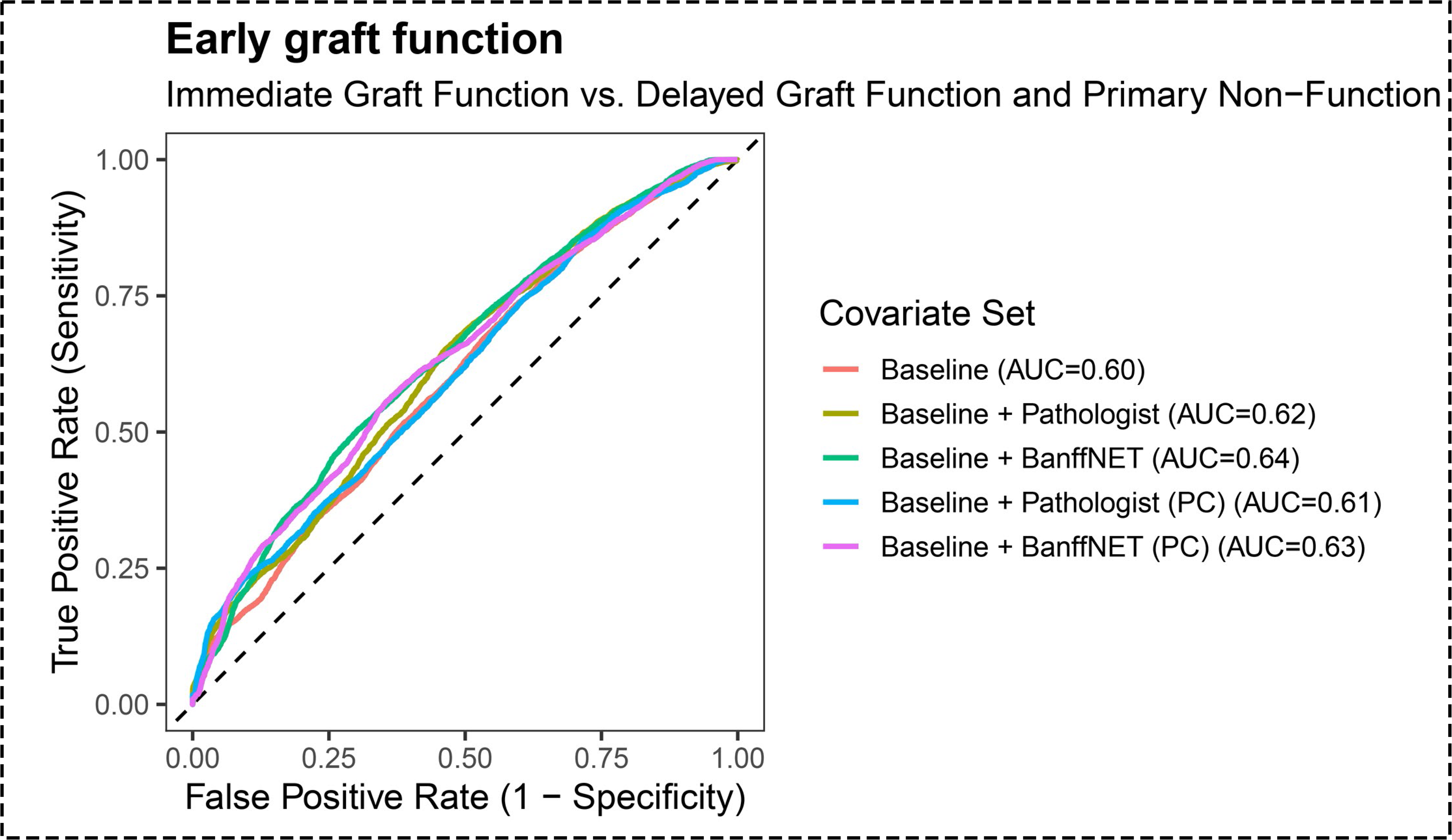
ROC curves of binary logistic regression models using five covariate sets. Each colored line represents a model with a different covariate set. Binary classification distinguished IGF from non-IGF (DGF and PNF combined). Model performance was evaluated using the area under the receiver operating characteristic curve (AUC)

Given the small number of PNF cases (n = 19), a penalized multinomial ridge regression model was fitted as a sensitivity analysis. Associations are presented in Supplementary Figure 5, and ROC analyses are shown in Figure 3. As summarized in Table 3, extending the baseline clinical model with pathologist-assessed scores resulted in minimal changes in performance (Δaccuracy = +0.01; ΔAUC = 0.00). Similarly, adding the PCs of pathologist-assessed scores did not improve model performance (Δaccuracy = −0.01; ΔAUC = 0.01). In contrast, extending the baseline clinical model with BanffNET scores improved accuracy by 0.03 and AUC by 0.03, with similar gains observed when incorporating the PCs of BanffNET features (Δaccuracy = +0.04; ΔAUC = +0.04).

**Figure 3.**
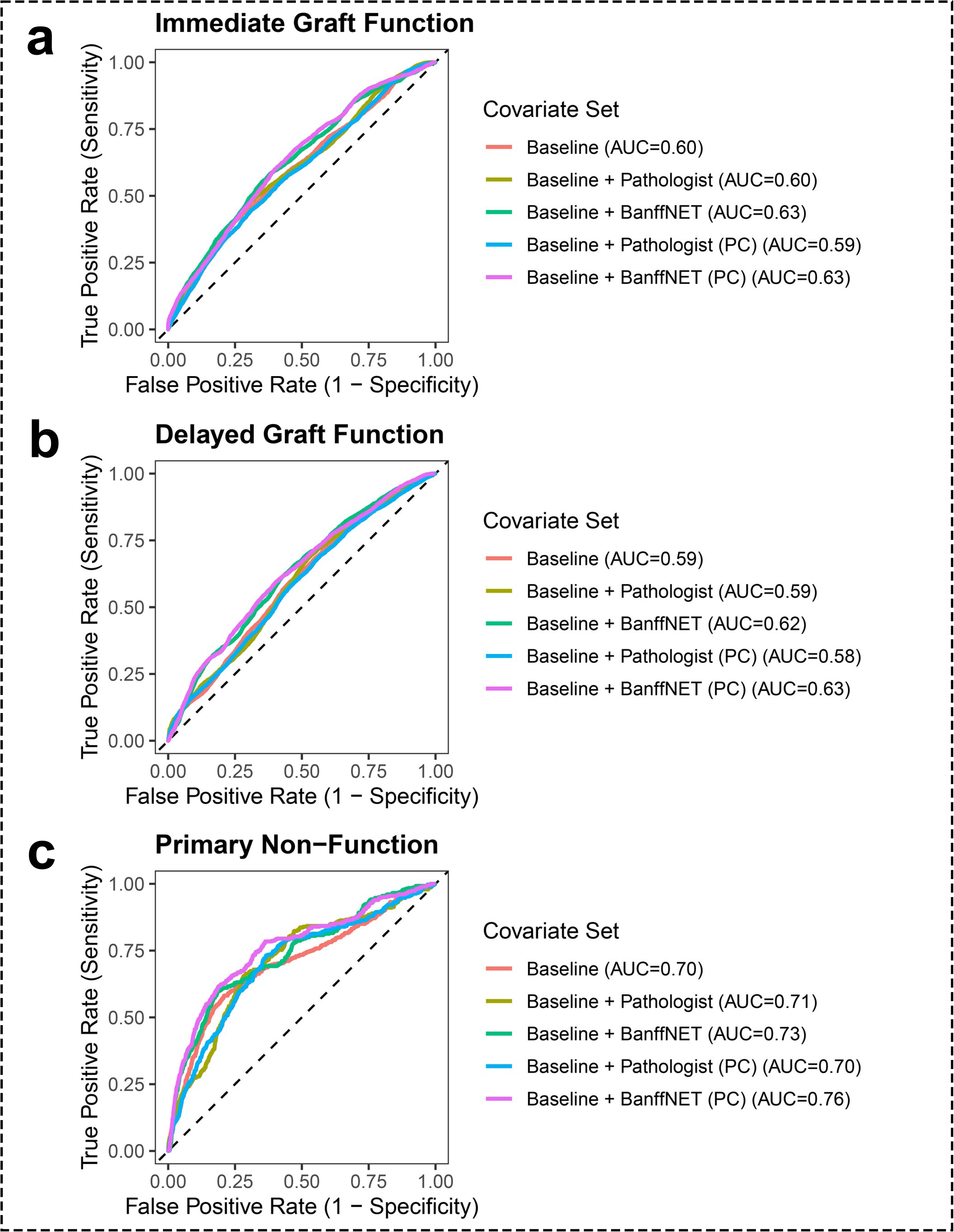
ROC curves of penalized multinomial regression models using five covariate sets. Discriminative performance was evaluated for three one-vs-rest comparisons: IGF, DGF, and PNF. For each comparison, five models incorporating different covariate sets are shown (colored lines). AUC values were used to assess model performance.

**Table 3.** Penalized multinomial regression analysis for early graft function, model performance comparison.

| Comparison | Extended variables | Δ Accuracy | Δ AUC |
| --- | --- | --- | --- |
| Set 2 vs Set 1 | Pathologist scores | 0.01 | 0.00 |
| Set 3 vs Set 1 | BanffNET scores | 0.03 | 0.03 |
| Set 4 vs Set 1 | PC of pathologist scores | -0.01 | 0.01 |
| Set 5 vs Set 1 | PC of BanffNET scores | 0.04 | 0.04 |

AUC, area under the receiver operating characteristic curve; PC, principal component. Δ indicates the change compared with the baseline model (Set 1). Set 1 represents the Baseline model, while Sets 2–5 include additional covariates as specified.

### Incremental value of histopathological features in predicting kidney function trajectories

Compared with the baseline clinical model, the addition of histopathological features significantly improved model fit for both eGFR and UPCR trajectories across all comparisons, as indicated by LRT p-values < 0.05 and supported by negative ΔAIC values (Tables 4 and 5). However, the incremental contribution of histopathological features to explained variance was limited, with Δ marginal R² ranging from approximately 1% to 2% for both outcomes. Consistent with these findings, observed versus predicted trajectories for eGFR and UPCR (Figure 4a–b) showed that models incorporating histopathological features closely captured the longitudinal trends over time but resulted in only subtle changes compared with the baseline clinical model.

**Figure 4.**
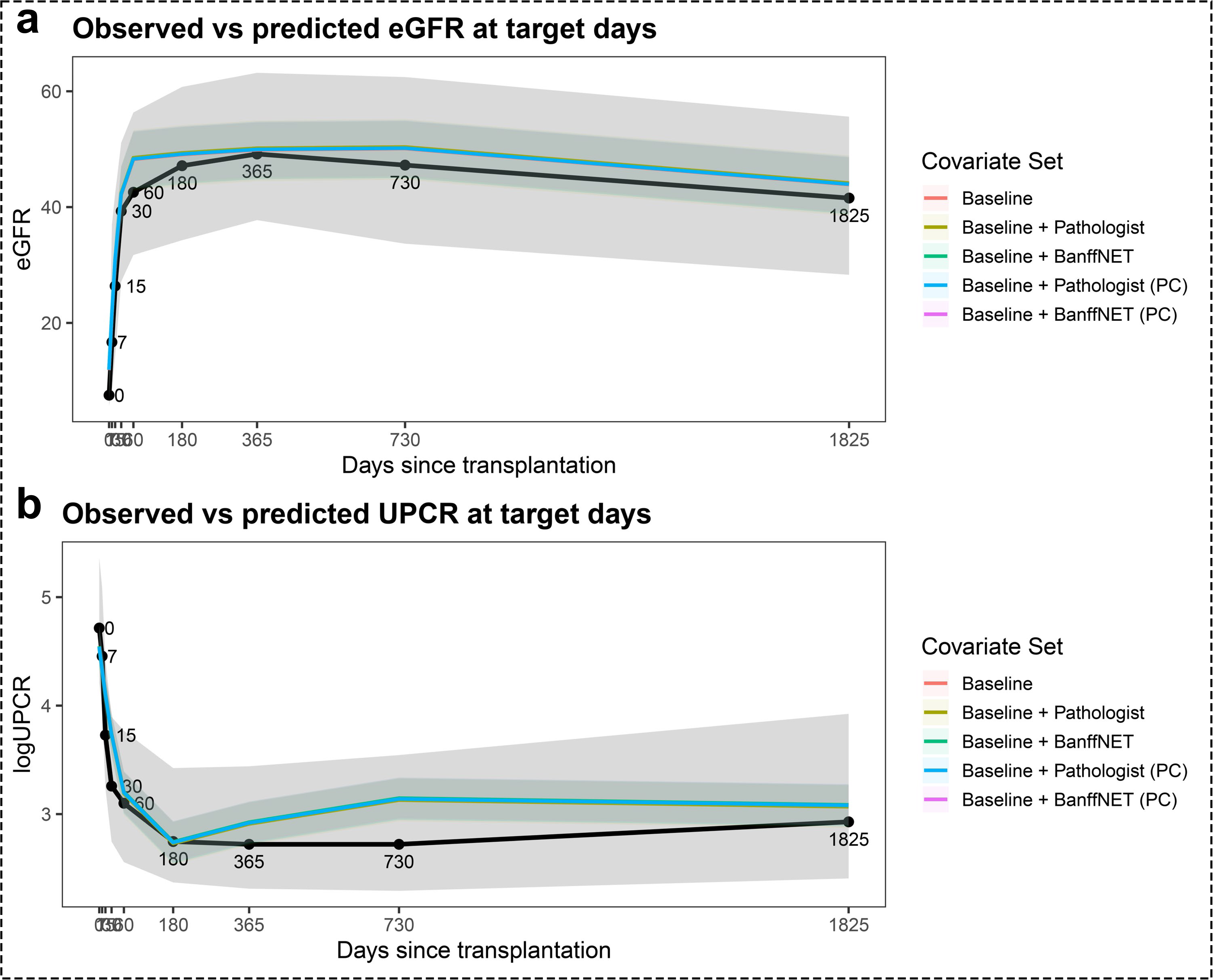
Observed vs. predicted kidney function trajectories (a: eGFR, b: UPCR) at predefined time points. Observed and model-predicted eGFR and logUPCR values are shown at target days 0, 7, 15, 30, 60, 180, 365, 730, and 1825. The black line represents observed values, summarized as the median across patients, with measurements assigned to target days using nearest-neighbor matching within predefined time windows. Colored lines represent predicted values from five models constructed using different covariate sets. Predictions were obtained from mixed-effects models and evaluated at exact target days using fixed effects only. Both observed and predicted summaries were pooled across multiple imputed datasets and are presented as medians (with interquartile ranges, if shown).

**Table 4.** Linear mixed model for eGFR trajectory, model performance comparison.

| Comparison | Extended variables | P value (LRT test) | $\Delta$ AIC | $\Delta$ marginal $R^2$ |
| --- | --- | --- | --- | --- |
| Set 2 vs Set 1 | Pathologist scores | 0.00 | -28.90 | 0.02 |
| Set 3 vs Set 1 | BanffNET scores | 0.04 | 5.21 | 0.01 |
| Set 4 vs Set 1 | PC of pathologist scores | 0.00 | -16.90 | 0.01 |
| Set 5 vs Set 1 | PC of BanffNET scores | 0.01 | -7.21 | 0.01 |

PC, principal component. LRT, likelihood ratio test; AIC, Akaike Information Criterion; marginal $R^2$ , variance explained by fixed effects; $\Delta$ indicates change compared with the baseline model (Set 1). Set 1 represents the baseline model, while Sets 2–5 include additional biopsy-derived variables as specified.

**Table 5.** Linear mixed effect model for UPCR trajectory, model performance comparison.

| Comparison | Extended variables | P value (LRT test) | $\Delta$ AIC | $\Delta$ marginal $R^2$ |
| --- | --- | --- | --- | --- |
| Set 2 vs Set 1 | Pathologist scores | 0.00 | -22.06 | 0.02 |
| Set 3 vs Set 1 | BanffNET scores | 0.01 | -0.77 | 0.02 |
| Set 4 vs Set 1 | PC of pathologist scores | 0.00 | -10.44 | 0.01 |
| Set 5 vs Set 1 | PC of BanffNET scores | 0.00 | -9.98 | 0.01 |

PC, principal component. LRT, likelihood ratio test; AIC, Akaike Information Criterion; marginal $R^2$ , variance explained by fixed effects; $\Delta$ indicates change compared with the baseline model (Set 1). Set 1 represents the baseline model, while Sets 2–5 include additional biopsy-derived variables as specified.

### Incremental value of histopathological features in predicting the need for a post-transplant indication biopsy

For both eGFR- and UPCR-based joint models predicting the need for a post-transplant indication biopsy, the baseline clinical model consistently demonstrated the best overall fit, as indicated by the lowest DIC and WAIC and the highest LPML compared with models augmented with additional histopathological features (Tables 6–7).

**Table 6.** Comparison of joint models (LME: eGFR trajectory, Cox: indication biopsy) across covariate sets based on model fit and model evidence.

| Covariates sets | Description | WAIC | DIC | LPML |
| --- | --- | --- | --- | --- |
| Set 1 | Baseline* | 10,215 | 10,256 | -5,108 |
| Set 2 | Baseline + Pathologist | 10,248 | 10,311 | -5,163 |
| Set 3 | Baseline + BanffNET | 10,318 | 10,389 | -5,221 |
| Set 4 | Baseline + Pathologist (PC) | 10,280 | 10,286 | -5,169 |
| Set 5 | Baseline + BanffNET (PC) | 10,243 | 10,301 | -5,139 |

\*Baseline, routine clinical characteristics; LME, linear mixed-effects model; Cox, Cox proportional hazards model; PATH, pathologist scores; PATH (PCA), principal components derived from pathologist scores; BanffNET (PCA), principal components derived from BanffNET scores; DIC, Deviance Information Criterion; WAIC, Watanabe–Akaike information criterion; LPML, log pseudo marginal likelihood.

**Table 7.** Comparison of joint models (LME: UPCR trajectory, Cox: indication biopsy) across covariate sets based on model fit and model evidence.

| Covariates sets | Description | WAIC | DIC | LPML |
| --- | --- | --- | --- | --- |
| Set 1 | Baseline* | 12,402 | 12,440 | -6,201 |
| Set 2 | Baseline + Pathologist | 12,427 | 12,486 | -6,242 |
| Set 3 | Baseline + BanffNET | 12,482 | 12,540 | -6,294 |
| Set 4 | Baseline + Pathologist (PC) | 12,424 | 12,466 | -6,239 |
| Set 5 | Baseline + BanffNET (PC) | 12,430 | 12,470 | -6,243 |

\*Baseline, routine clinical characteristics; LME, linear mixed-effects model; Cox, Cox proportional hazards model; PATH, pathologist scores; PATH (PCA), principal components derived from pathologist scores; BanffNET (PCA), principal components derived from BanffNET scores; DIC, Deviance Information Criterion; WAIC, Watanabe–Akaike information criterion; LPML, log pseudo marginal likelihood.

Time-dependent ROC analyses at 60-day landmark times across 10-year prediction horizons demonstrated discriminatory performance for prediction of post-transplant indication biopsy need using both eGFR and UPCR biomarkers, with cumulative dynamic AUC values ranging from 0.61 to 0.71. For eGFR-based models, the AUC values increased slightly during the early prediction horizons, peaking around years 2–3 (0.67-0.71), followed by a gradual decline toward years 8–10 (0.63-0.67) (Figure 5a). Similar patterns were observed for UPCR- based models, with the AUCs ranging from 0.61–0.71 across prediction horizons (Figure 5b). Models incorporating histopathological features consistently outperformed the baseline clinical model, but the absolute improvement in AUC was limited (ΔAUC 0.00 to 0.06.

**Figure 5.**
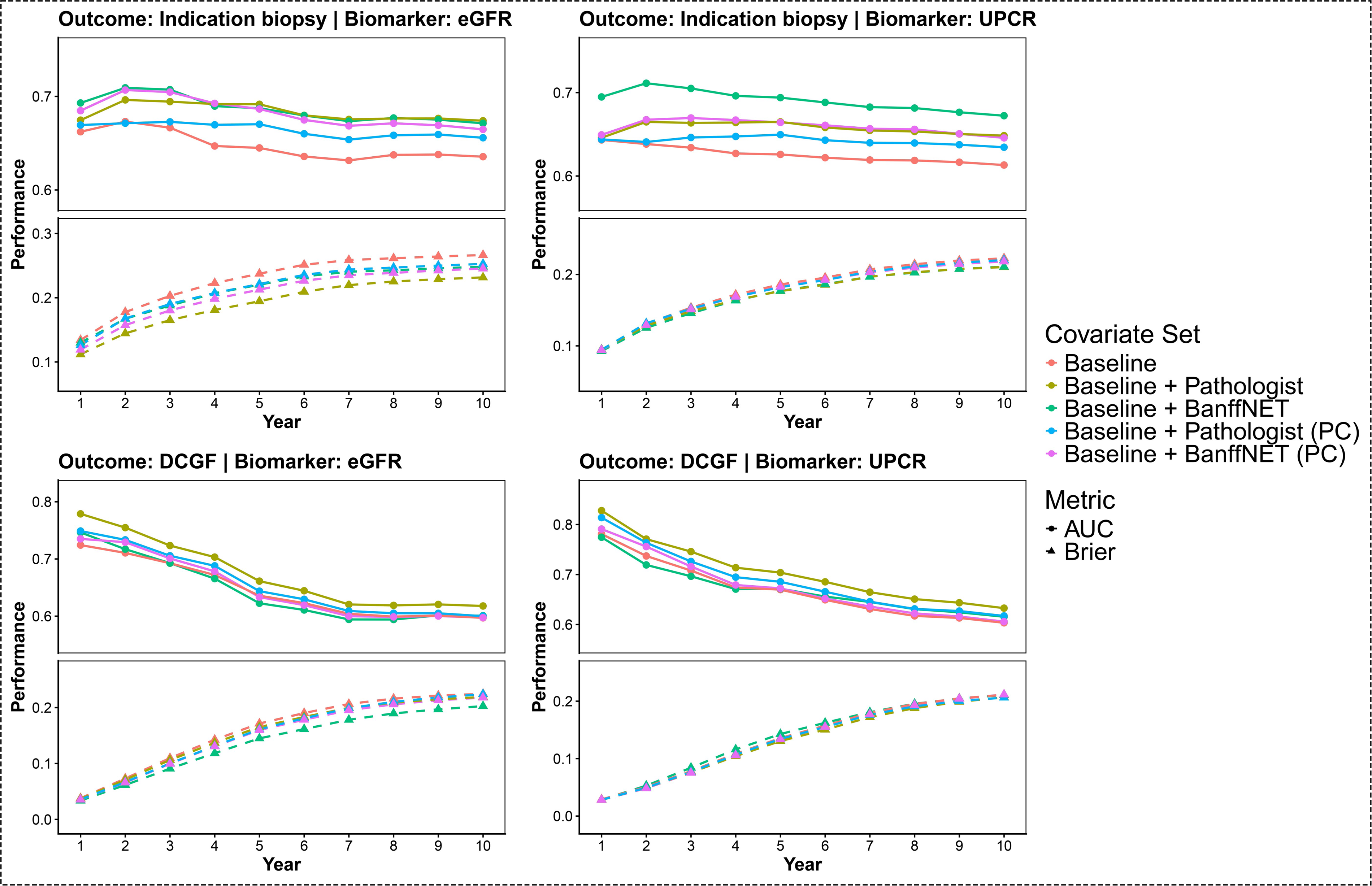
Dynamic prediction performance across covariate sets at the 60-day landmark over 10-year prediction horizons. Panels (a–b) show prediction performance for indication biopsy, and panels (c–d) show prediction performance for death-censored graft failure (DCGF). Models were evaluated using eGFR (a, c) and UPCR (b, d) as longitudinal biomarkers. Solid circles represent time-dependent area under the receiver operating characteristic curve (AUC), and dashed triangles represent Brier scores across 10-year prediction horizons. Covariate sets included baseline clinical variables alone, baseline plus BanffNET features, baseline plus BanffNET principal components (PC), baseline plus pathologist scores(PATH), and baseline plus PATH PC.

Models that included PCs of BanffNET scores generally demonstrated the highest discrimination performance for UPCR-based prediction, with mean ΔAUC 0.06, whereas other models showed mean ΔAUC improvements of 0.02–0.03 (Supplementary table 2)

Prediction error increased over longer prediction horizons for both biomarkers, as reflected by gradually increasing Brier scores. In eGFR-based models, Brier scores increased from 0.11–0.13 at early prediction horizons to 0.23-0.27 by year 10 (Figure 5a). Similarly, in UPCR-based models, Brier scores increased from approximately 0.09 at early horizons to 0.22 during follow-up (Figure 5b). Models incorporating histopathological features consistently demonstrated lower prediction error than the baseline clinical model alone, as indicated by negative ΔBrier scores (Supplementary Table 3).

### Incremental value of histopathological features in predicting death-censored graft failure

For both eGFR- and UPCR-based joint models predicting DCGF, the baseline clinical model (Set 1) provided the best overall performance, yielding the lowest DIC and WAIC values and the highest LPML relative to models additionally incorporating histopathological features (Sets 2–5) (Tables 8–9).

**Table 8.** Comparison of joint models (LME: eGFR trajectory, COX: DCGF) across covariate sets based on model fit and model evidence.

| Covariates sets | Description | WAIC | DIC | LPML |
| --- | --- | --- | --- | --- |
| Set 1 | Baseline* | 9,901 | 9,940 | -4,954 |
| Set 2 | Baseline + Pathologist | 9,957 | 10,015 | -5,221 |
| Set 3 | Baseline + BanffNET | 10,058 | 10,096 | -5,535 |
| Set 4 | Baseline + Pathologist (PC) | 9,983 | 9,984 | -5,123 |
| Set 5 | Baseline + BanffNET (PC) | 9,931 | 9,990 | -5,031 |

\*Baseline, routine clinical characteristics; LME, linear mixed-effects model; Cox, Cox proportional hazards model; PATH, pathologist scores; PATH (PCA), principal components derived from pathologist scores; BanffNET (PCA), principal components derived from BanffNET scores; DIC, Deviance Information Criterion; WAIC, Watanabe–Akaike information criterion; LPML, log pseudo marginal likelihood.

**Table 9.**
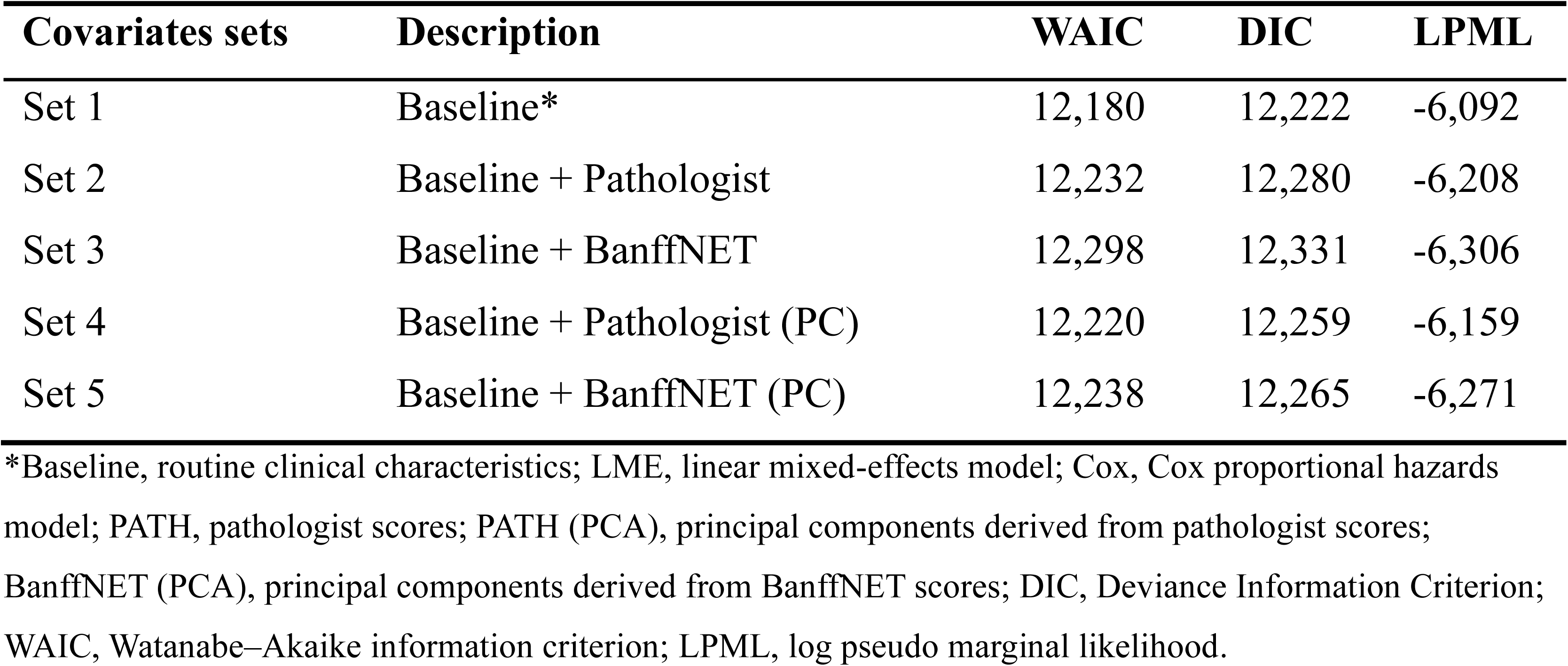
Comparison of joint models (LME: UPCR trajectory, Cox: DCGF) across covariate sets based on model fit and model evidence.

| Covariates sets | Description | WAIC | DIC | LPML |
| --- | --- | --- | --- | --- |
| Set 1 | Baseline* | 12,180 | 12,222 | -6,092 |
| Set 2 | Baseline + Pathologist | 12,232 | 12,280 | -6,208 |
| Set 3 | Baseline + BanffNET | 12,298 | 12,331 | -6,306 |
| Set 4 | Baseline + Pathologist (PC) | 12,220 | 12,259 | -6,159 |
| Set 5 | Baseline + BanffNET (PC) | 12,238 | 12,265 | -6,271 |

\*Baseline, routine clinical characteristics; LME, linear mixed-effects model; Cox, Cox proportional hazards model; PATH, pathologist scores; PATH (PCA), principal components derived from pathologist scores; BanffNET (PCA), principal components derived from BanffNET scores; DIC, Deviance Information Criterion; WAIC, Watanabe–Akaike information criterion; LPML, log pseudo marginal likelihood.

Time-dependent ROC analyses using 60-day landmark times across 10-year prediction horizons for DCGF showed progressively declining discrimination with increasing prediction horizon length for both eGFR- and UPCR-based models. For eGFR-based prediction, AUCs decreased from approximately 0.72–0.78 at year 1 to 0.60–0.62 by year 10 (Figure 5c).

Similarly, UPCR-based models declined from approximately 0.79–0.83 at year 1 to 0.60–0.62 at year 10 (Figure 5d). Across prediction horizons, models incorporating histopathological features consistently showed slightly higher discrimination than the baseline clinical model, although the improvements were limited, with mean ΔAUC values ranging from 0.00 to 0.04. The largest gains were observed for models incorporating pathologist-assessed scores, with mean ΔAUC improvements of 0.03 for eGFR-based prediction and 0.04 for UPCR-based prediction (Supplementary Table 2).

Brier scores increased steadily over longer prediction horizons for both biomarkers, rising from approximately 0.03–0.04 at year 1 to 0.20–0.22 by year 10, indicating reduced predictive accuracy over time (Figure 5c–d). Differences between models incorporating histopathological features and the baseline clinical model remained small, with ΔBrier values ranging from −0.02 to 0.00 (Supplementary Table 3). Overall, these findings indicate that histopathological features provided only limited direct incremental predictive value beyond baseline clinical characteristics.

### The continuous BanffNET score associates with early and late graft outcomes

The Banff Time-Zero working group suggested that the pre-implantation biopsy could potentially salvage donor kidneys that were considered untransplantable based in clinical data only. The role of the kidney biopsy would be to rule out chronic damage. To evaluate risk across the full spectrum of donor quality and histopathology, we mapped continuous composite BanffNET scores against continuous KDRI. For early graft dysfunction (Non-IGF), the bivariate surface demonstrated a progressive risk gradient (Figure 6a-b).

**Figure 6.**
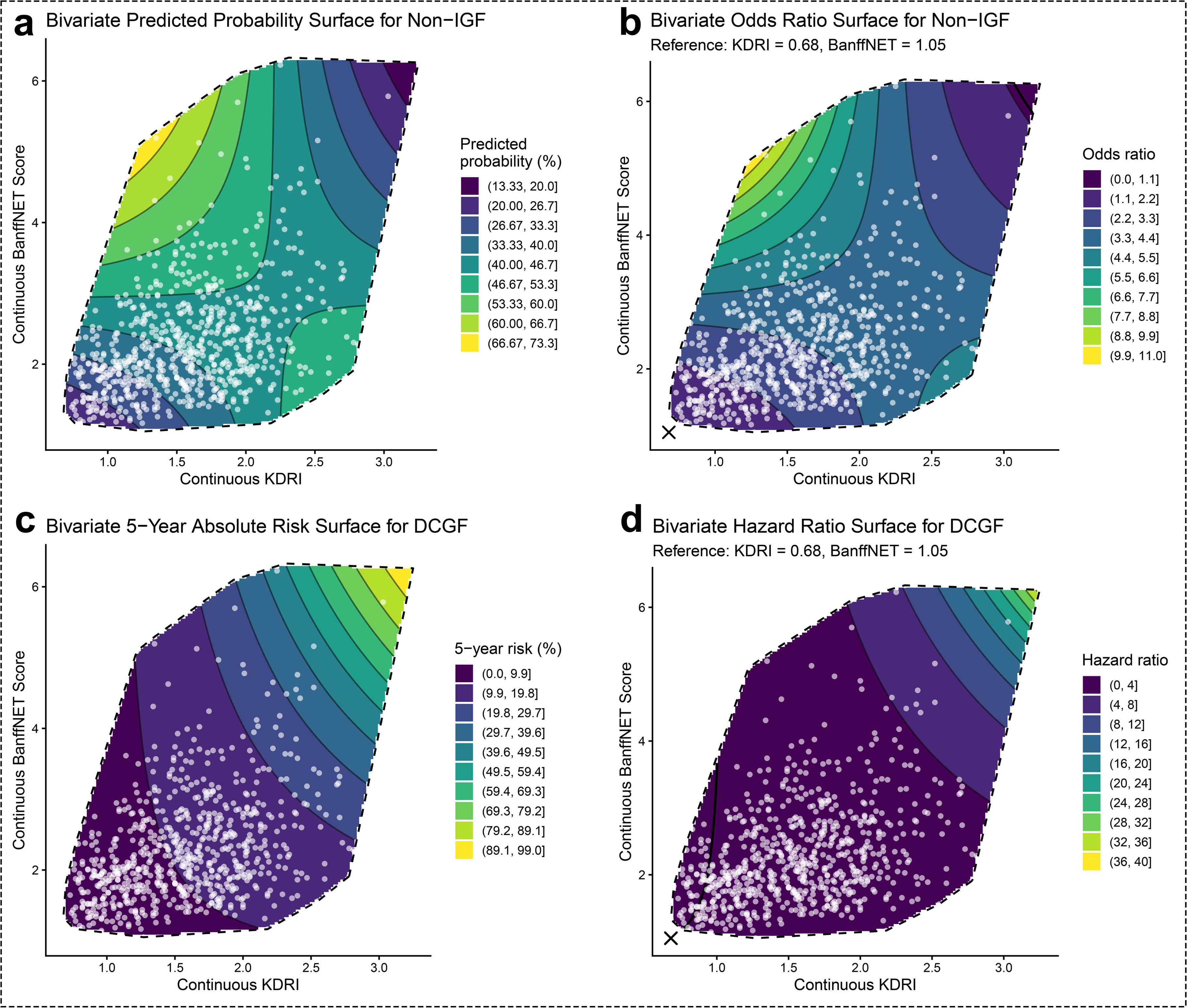
Bivariate associations of KDRI and composite BanffNET scores with non- immediate graft function (non-IGF) and death-censored graft failure (DCGF). Bivariate surface plots illustrate the joint associations of KDRI (x-axis) and BanffNET score (y-axis) with transplant outcomes. (a) Predicted probability of non-IGF. (b) Odds ratio for non-IGF, relative to KDRI = 0.68 and BanffNET score = 1.05. (c) Predicted 5-year absolute risk of DCGF. (d) Hazard ratio for DCGF, relative to KDRI = 0.68 and BanffNET score = 1.05. Colored surfaces represent the magnitude of the predicted probability, odds ratio, absolute risk, or hazard ratio according to the corresponding color scales, with contour lines indicating levels of equal predicted outcome or relative effect. White points show the observed distribution of KDRI and BanffNET score in the study population. Dashed boundaries delineate the region of the covariate space supported by the observed data.

Interestingly, the predicted probability of Non-IGF scaled up to 73.3% in the extreme upper-left quadrant, representing low KDRI but maximum composite BanffNET scores. Correspondingly, the odds ratio for Non-IGF peaked in this discordant high-risk continuum compared to the reference means.

For DCGF, the continuous composite surfaces revealed a different synergistic risk topography. The 5-year absolute risk of DCGF approached 99.0% at the highest combined extremes of both KDRI and BanffNET (the upper-right quadrant), which corresponded to hazard ratios exceeding 29.7 relative to the reference means (Figure 6c-d).

Deconstructing the composite score into individual continuous lesions revealed heterogeneity in risk topologies. Continuous evaluations of specific lesions, such as vascular changes (ah, cv) and chronicity indices (ifta, iifta), plotted against KDRI demonstrated distinct non-linear risk gradients for early dysfunction probabilities and odds ratios (Supplementary Figure 6a- b), as well as DCGF absolute risks and hazard ratios (Supplementary Figure 7a-b). This indicates that specific histological phenotypes interact differently with clinical donor risk profiles across their continuous spectra, rather than acting as uniform additive penalties.

## Discussion

This study investigated the prognostic value of pre-implantation biopsy histopathology, particularly beyond known baseline donor and recipient characteristics risk factors for adverse outcome in the context of kidney transplant allocation. Histology was assessed using an optimal laboratory methodology combining conventional triple staining, expert renal pathologist evaluation, and the automated Banff lesion quantification algorithm (BanffNET). Our comparison of clinical-only models versus combined clinical-histological models revealed that histological features in general, whether assessed by a renal pathologist or by quantitative deep learning, provided only marginal improvements in predictive performance. Corroborating on the existing literature, we therefore question the clinical utility for the use of pre-implantation in the setting of organ allocation.

Several studies have reported the prognostic value of pre-implantation biopsy; however, the evidence remains insufficiently robust. Variability in sample taking, processing and evaluation can substantially reduce reproducibility, and an unreliable histopathological ground truth may undermine the validity and confidence of subsequent analyses. To overcome some of these challenges, recent studies have applied deep learning models to improve the consistency and objectivity of biopsy assessment, including models based on frozen-section procurement biopsies for predicting graft outcomes^27^, AI-assisted tools for supporting allocation decisions^28,29^, and approaches focusing on global histological damage or isolated features such as glomerulosclerosis^30^. While these efforts demonstrate the potential of AI in this setting, they remain constrained by limitations in tissue quality, coarse-grained or feature-restricted representations, and reliance on suboptimal scoring frameworks. In this context, our study addresses these limitations by integrating optimal histopathological conditions with a computational pipeline-assisted, enabling lesion-specific and continuous evaluation for a more precise and standardized characterization of histopathological features.

Beyond optimizing histopathological assessment, another key strength of this study lies in the statistical framework used to evaluate the contribution of histopathological features. Although prior studies have explored the use of pre-implantation biopsy findings for predictive modeling, this was not our primary objective. Instead, we aimed to quantify the incremental value of histopathological features beyond baseline clinical characteristics using a range of complementary statistical approaches. Importantly, recent studies have largely relied on summary discrimination metrics such as the C-index and AUC^29,31,32^, while overlooking other critical aspects, including reductions in prediction error, improvements in model fit, and whether the added complexity of biopsy-based parameters is justified. In contrast, our study assessed model performance across multiple dimensions, providing a more comprehensive evaluation of the role of pre-implantation biopsy and enabling a balanced assessment of its potential clinical utility in kidney transplant allocation.

Notably, even under these optimized conditions, the added prognostic value of histopathological features remained limited, suggesting that their role in kidney transplant allocation may be inherently constrained when also considering clinical parameters. First, transplant management is highly complex both before and after transplantation^33^, with numerous factors influencing graft outcomes that may attenuate the predictive contribution of pre-implantation biopsy findings. Prior to transplantation, extensive efforts are undertaken in donor–recipient matching and immunological risk assessment to reduce the likelihood of rejection^34^. Advanced perfusion techniques have also been proposed to reduce ischemia– reperfusion injury^9,35^, while evolving immunosuppressive regimens aim to balance rejection risk and treatment-related adverse effects^36^. Furthermore, post-transplant factors, particularly recipient adherence and self-management, represent substantial and often unpredictable influences on long-term graft survival^37^. It is important to mention that donor-derived chronic damage is not a dynamic risk factor that can be optimized, it represents the baseline loss of functional kidney parenchyma. Taken together, these considerations raise an important question: if routinely collected donor and recipient characteristics already capture nearly the maximum predictive value achievable under current, inherently constrained conditions for organ quality and graft survival, while biopsy findings offer only marginal incremental prognostic value, should biopsy-based information continue to play a decisive role in final allocation decisions?

The Banff Time-Zero Working Group recommended that based on available data, including data from the British randomized Pithia trial^38^, implantation biopsies in the context of organ allocation could best be used to salvage donor kidneys with a high risk of discard.

Incorporating continuous bivariate surface analysis demonstrates that the transition from acceptable to unacceptable risk is a topographical gradient rather than a binary step. Mapping continuous BanffNET scores against KDRI provides a highly granular framework for identifying viable “safe harbors” for organ salvage. Deconstructing these surfaces into individual continuous BanffNET lesions reveals distinct interactions with baseline clinical risk, which can be divided between established chronicity markers and novel inflammatory lesions. Quantitative BanffNET socres corresponding to risk factors routinely collected from pre-implantation biopsies (gs, ifta, cv, ah) display heterogeneous associations with short- and long-term outcomes. Interstitial fibrosis and tubular atrophy (ifta) and global glomerulosclerosis (gs) act as compounding risk multipliers for long-term graft failure primarily when superimposed on high-KDRI kidneys. Conversely, chronic vascular changes such as arteriolar hyalinosis (ah) and vascular fibrous intimal thickening (cv) exhibit broad risk gradients for early graft dysfunction that penalize even lower-KDRI organs. Beyond established chronicity, the continuous assessment of inflammatory and microvascular lesions uncovers unrecognized synergistic risks. Active chronic inflammation (iifta) creates steeper, more compressed risk gradients for long-term graft failure than inactive scarring (ifta alone). Furthermore, acute microvascular inflammation, such as peritubular capillaritis (ptc), albeit rare, displays dramatic risk peaks for early graft dysfunction even in clinically pristine, low- KDRI donor kidneys, while intimal arteritis (v) maintains concentrated hazard ratio peaks for long-term failure. Utilizing these continuous clinical-histological bivariate surfaces could potentially allow clinicians to pinpoint exact intersections where specific morphological phenotypes either safely permit transplantation or synergistically mandate discard.

A limitation of this study is that only wedge biopsies could be analysed, as this is the standard procedure in our hospital. Another limitation is that this is a single-center study. The findings would be more robust if corroborated in a multicenter setting. Secondly, also in our cohort we could not perform histological assessment of kidney donors discarded based on clinical decisions. This means that there is likely a bias towards the inclusion of lower KDRI donor kidneys. Most importantly, BanffNET is optimized for FFPE-derived digital whole slide images as this is the gold standard for optimal morphology. Ultrafast FFPE processing and staining would likely be required; morphological information is more difficult to pick up from frozen sections. This inherently prolongs cold ischemia time, although machine perfusion might mitigate the detrimental effects of extended preservation, safely widening the preservation window to allow for high-quality histological evaluation without compromising graft viability.

In conclusion, our study validate that overcoming technical and interpretive limitations still leaves pre-implantation biopsies with only marginal prognostic value over baseline donor and recipient clinical characteristics. This challenge their current clinical utility and highlight the urgent need to re-evaluate their use to prevent the unnecessary discard of potentially viable donor kidneys. Automated quantification with BanffNET based on digital whole slide images might further improve histological analysis of implantation biopsies to overcome interobserver variation and improve risk stratification of high risk donor kidneys.

## Disclosure statement

The authors declare that they have no competing interests relevant to this work.

## Data Sharing Statement

The data that support the findings of this study are available from the corresponding author upon reasonable request, subject to institutional, ethical and legal restrictions. A patent application (XXX) has been filed by XXX for the BanffNET deep learning algorism presented in this study.

## Data Availability

All data produced in the present study are available upon reasonable request to the authors

## Acknowledgments

The study is supported by the China Scholarship Council.

---

**Supplementary Figure 1.**
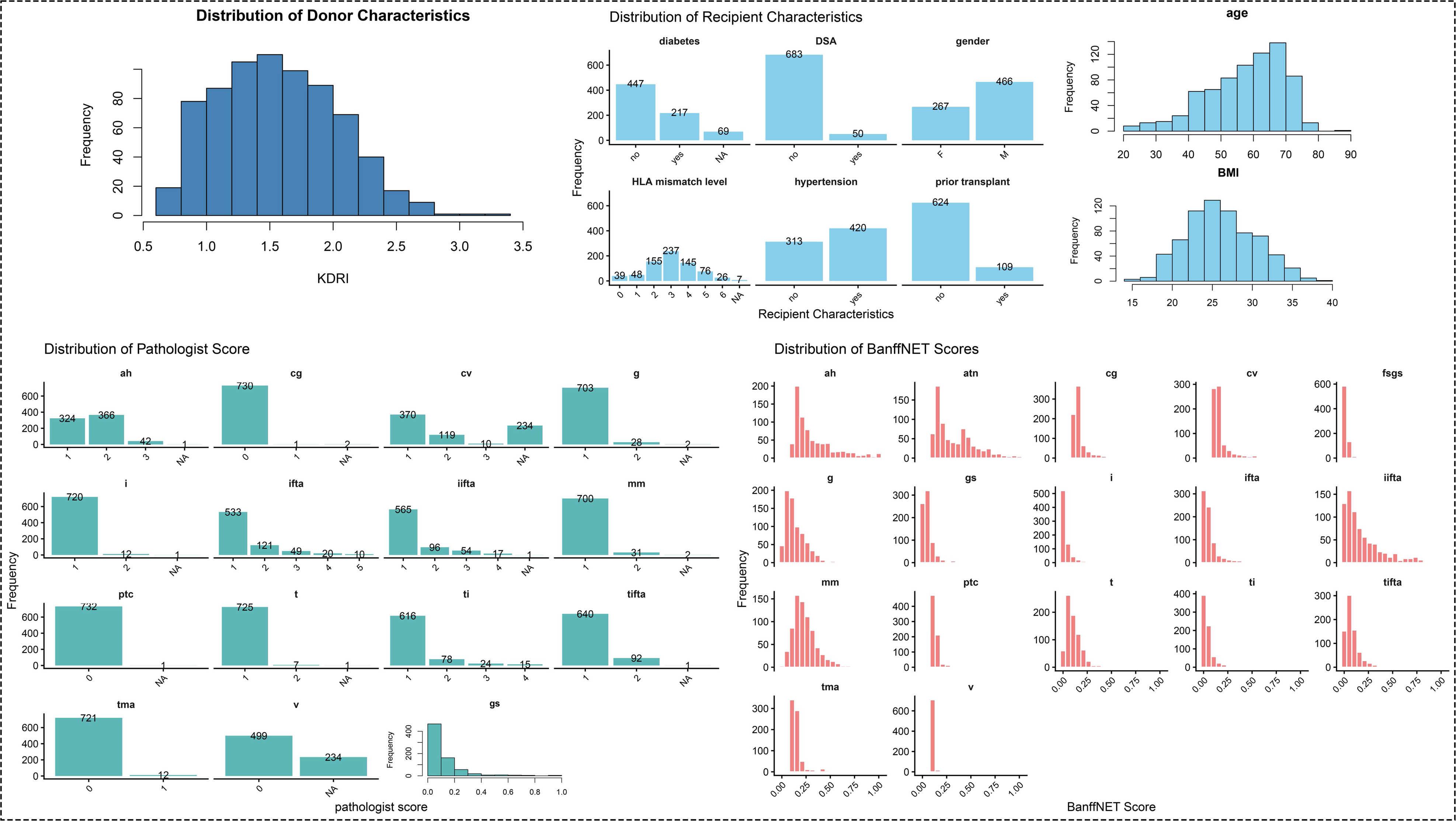
Distribution of KDRI, recipient characteristics, pathologist scores, and BanffNET scores. Histograms and bar charts show the distribution of all variables in the study cohort.

**Supplementary Figure 2.**
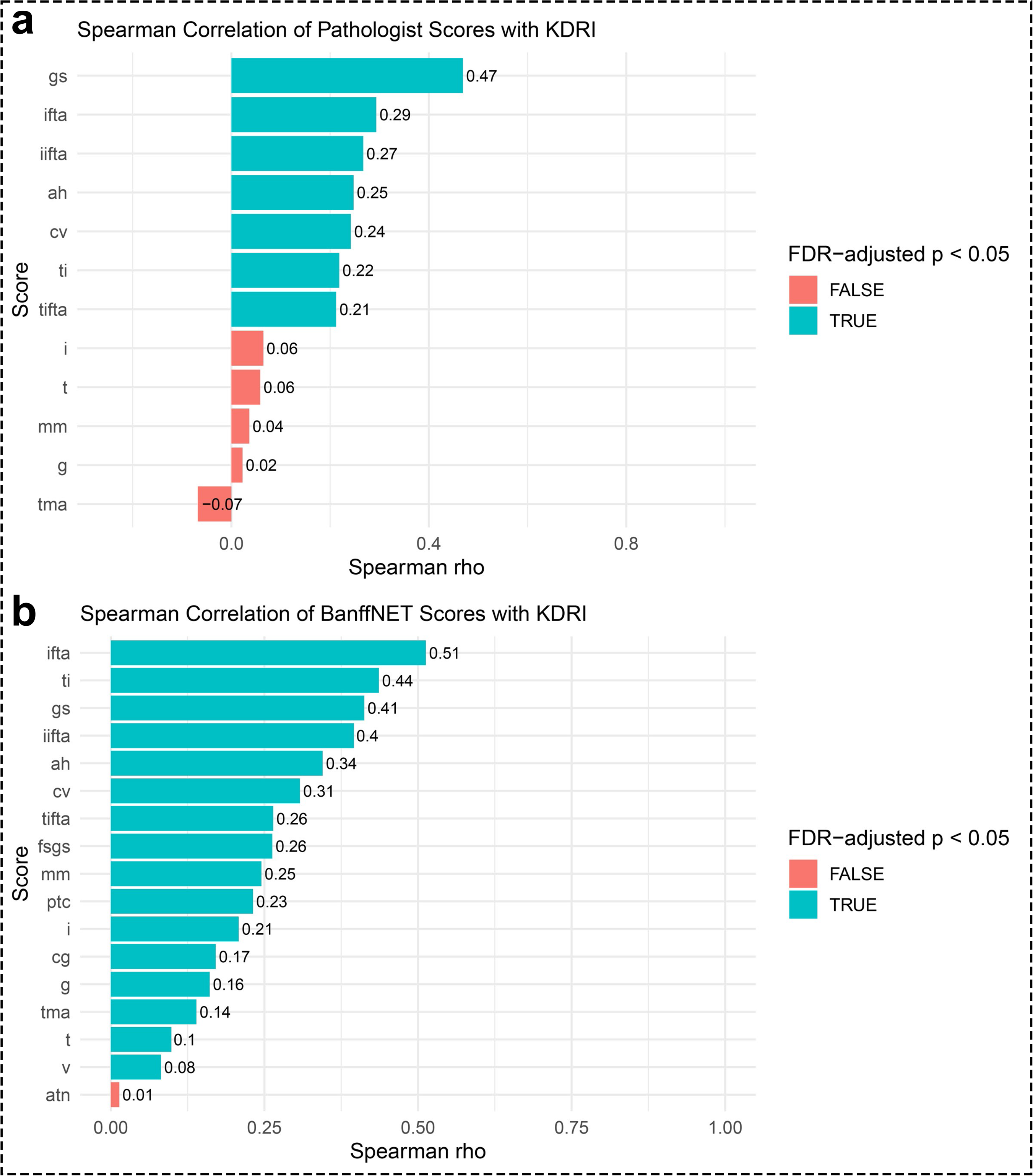
Spearman correlations between histopathological scores (a represents pathologist scores and b represents BanffNET scores ) and KDRI. Bar plot showing Spearman correlation coefficients (ρ) between KDRI and individual histopathological scores. Each bar represents the correlation for a specific score, with values displayed on the bars. Statistical significance was assessed using Spearman’s rank correlation test, and p-values were adjusted for multiple comparisons using the Benjamini–Hochberg false discovery rate (FDR) method. Bars are colored according to statistical significance (FDR-adjusted p < 0.05).

**Supplementary Figure 3.**
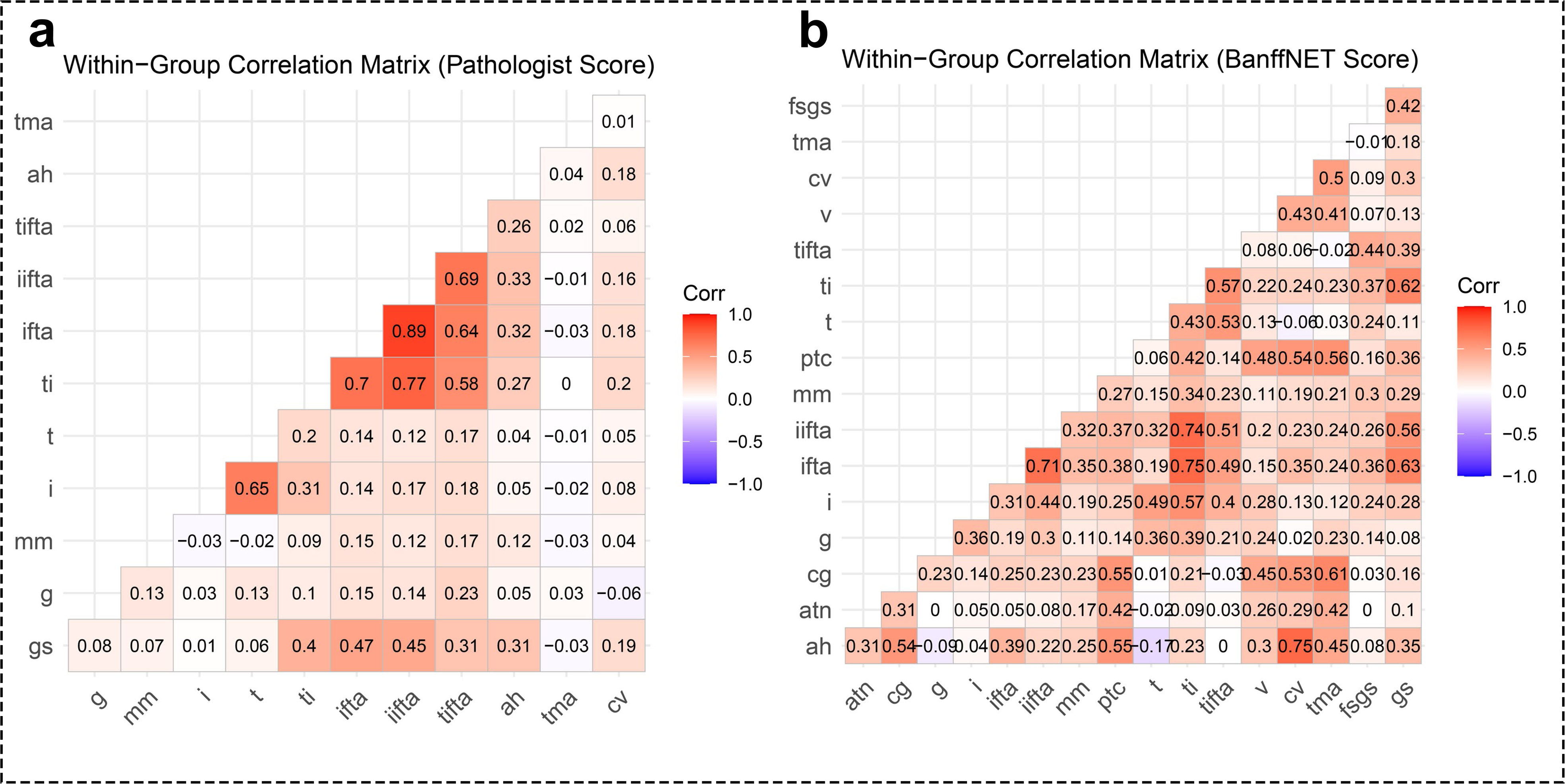
Correlation matrix of Pathologist scores (a) and BanffNET scores (b). Spearman correlation coefficients (ρ) were calculated between pathologist scores (BanffNET features) using pairwise complete data. The lower triangular matrix displays correlation values, summarizing interrelationships among histopathological scores.

**Supplementary Figure 4.**
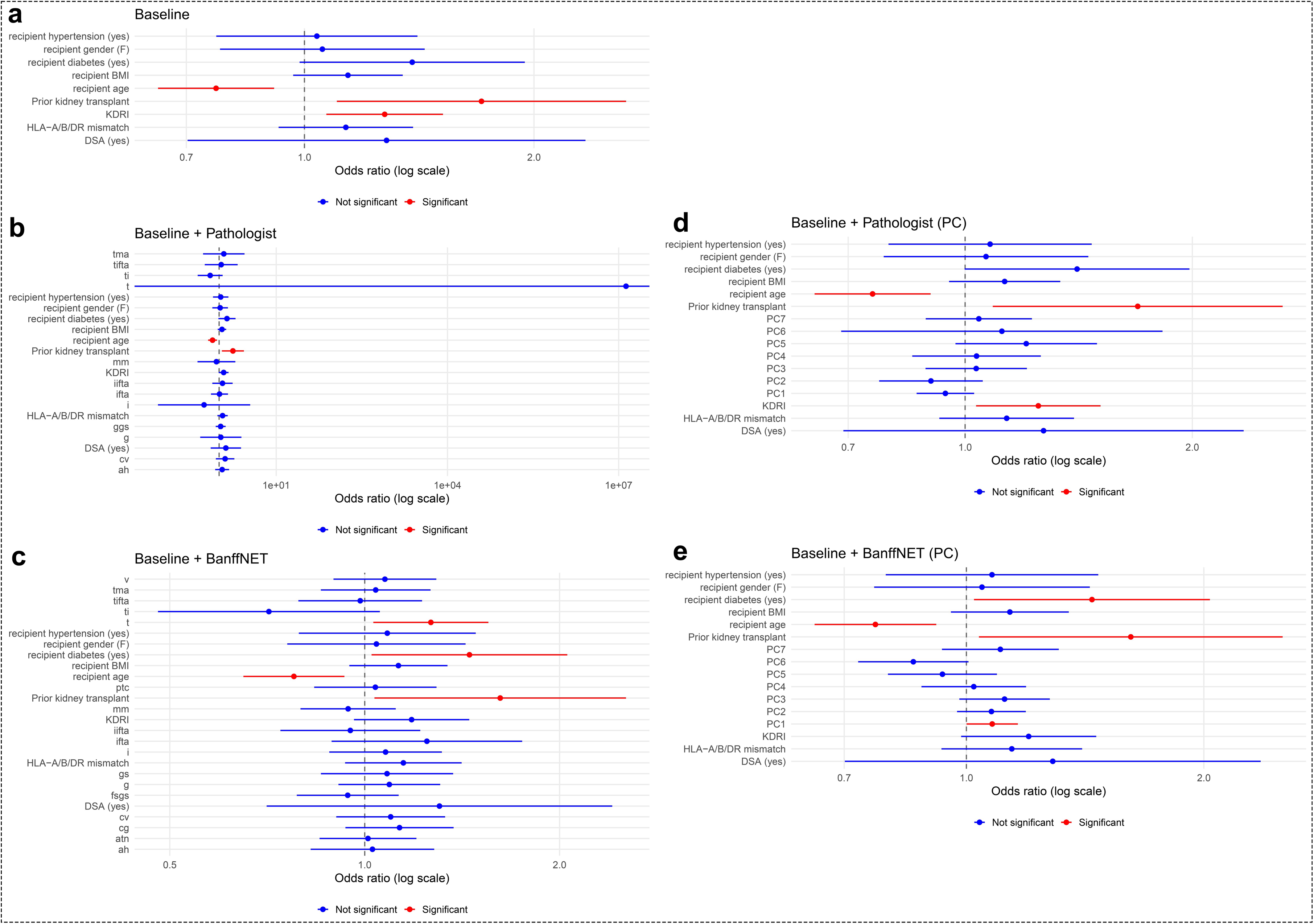
Forest plots of pooled odds ratios across five covariate sets. Forest plots showing pooled odds ratios (ORs) and 95% confidence intervals for predictors in each model: Baseline, Baseline + Pathologist Score, Baseline + BanffNET Score, Baseline + Pathologist Score (PCA), and Baseline + BanffNET Score (PCA). The dashed vertical line indicates OR = 1. Odds ratios are shown on a logarithmic scale. Colors indicate whether the 95% confidence interval excludes 1.

**Supplementary Figure 5.**
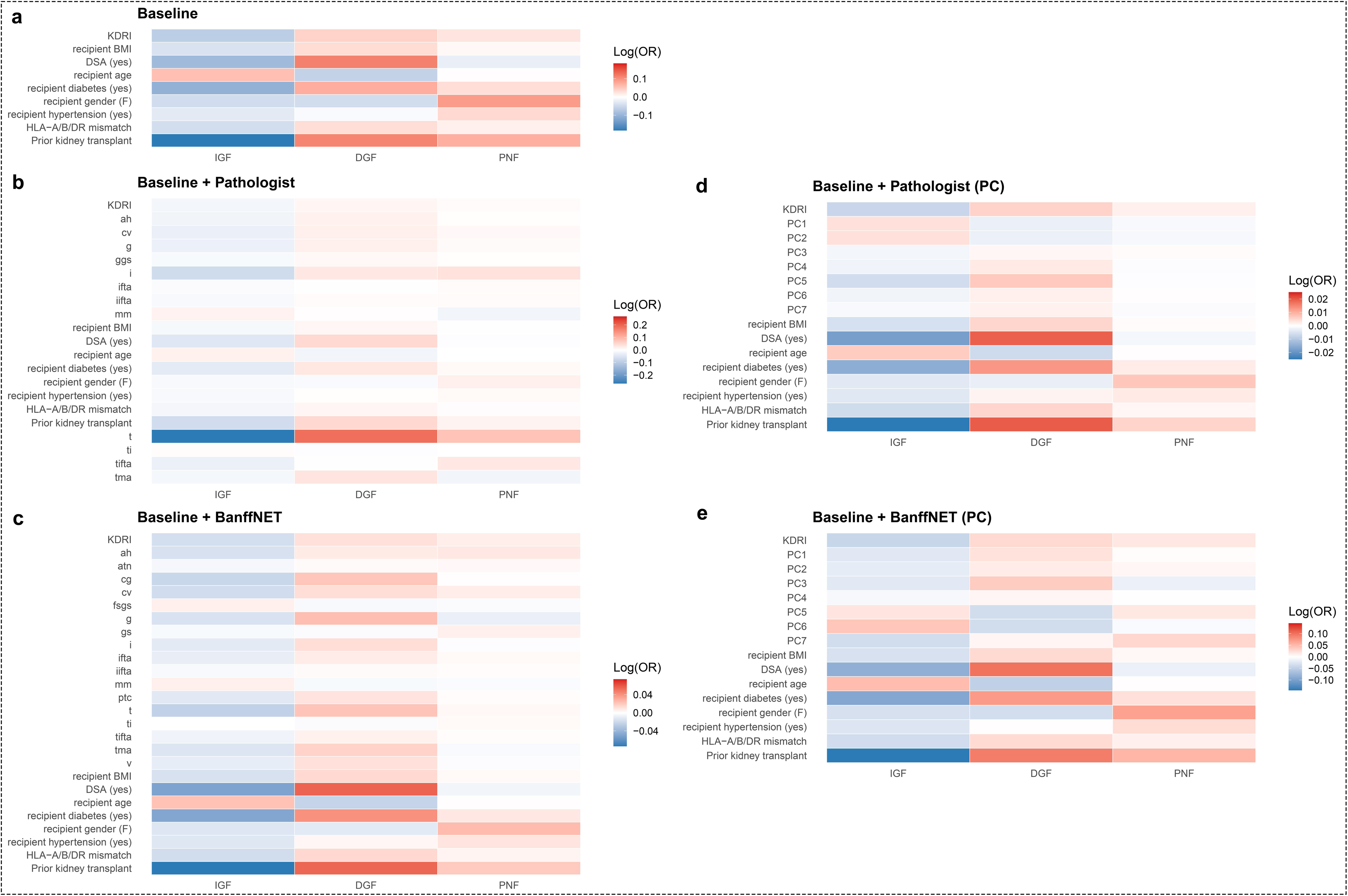
Heatmaps of penalized multinomial regression coefficients across five covariate sets. Heatmaps showing coefficient estimates (log odds ratios) from penalized multinomial regression models for predictors across outcome categories (IGF, DGF, PNF). Rows represent predictors and columns represent outcome categories. Color indicates the magnitude and direction of the coefficients (red: positive; blue: negative).

**Supplementary Figure 6.**
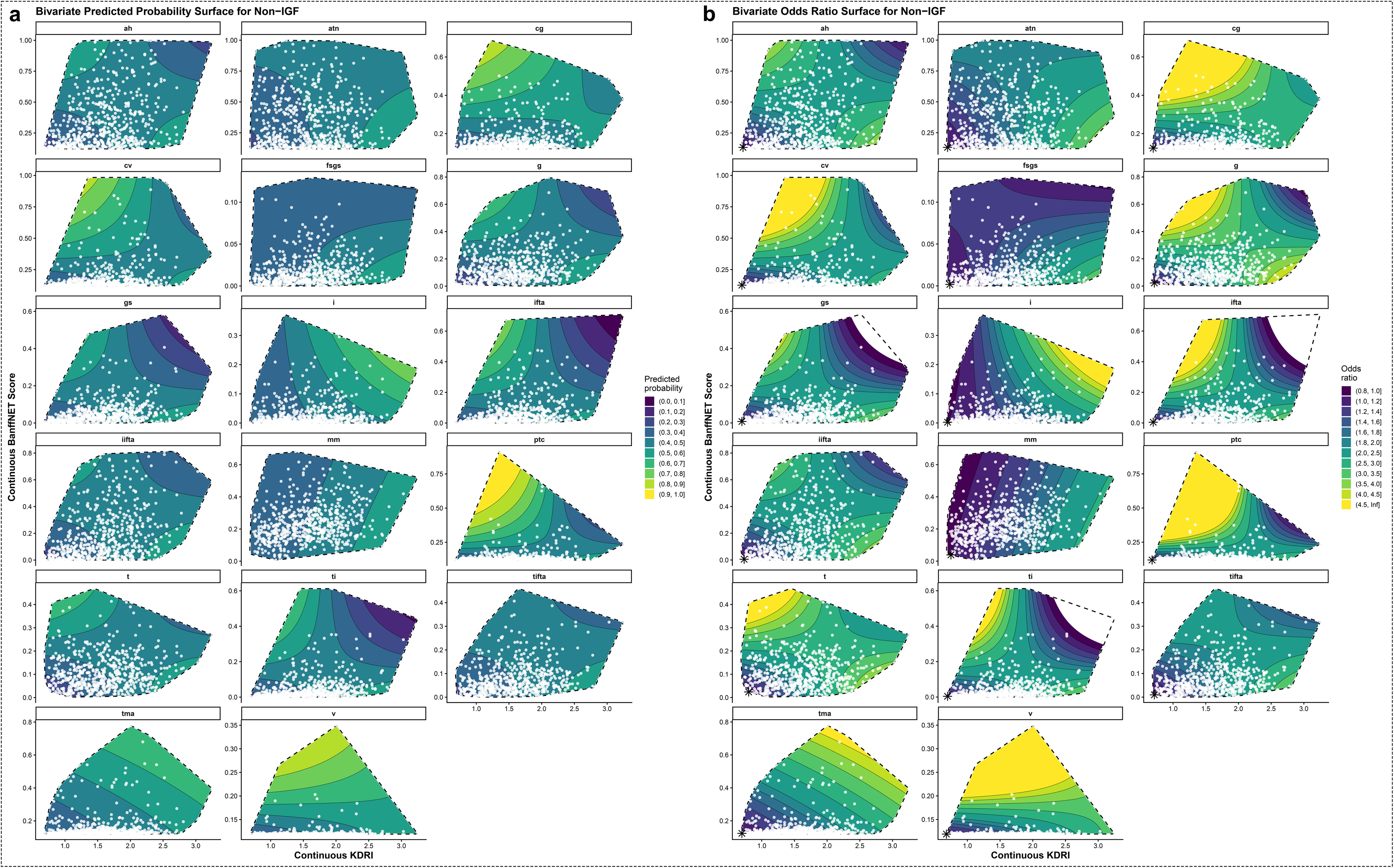
Bivariate associations of KDRI and individual BanffNET scores with non-immediate graft function (non-IGF). Bivariate surface plots illustrate the joint associations of KDRI (x-axis) and BanffNET score (y-axis) with transplant outcomes. (a) Predicted probability of non-IGF. (b) Odds ratio for non-IGF, relative to KDRI = 0.68 and BanffNET score = 1.05. Colored surfaces represent the magnitude of the predicted probability, odds ratio, absolute risk, or hazard ratio according to the corresponding color scales, with contour lines indicating levels of equal predicted outcome or relative effect. White points show the observed distribution of KDRI and BanffNET score in the study population. Dashed boundaries delineate the region of the covariate space supported by the observed data.

**Supplementary Figure 7.**
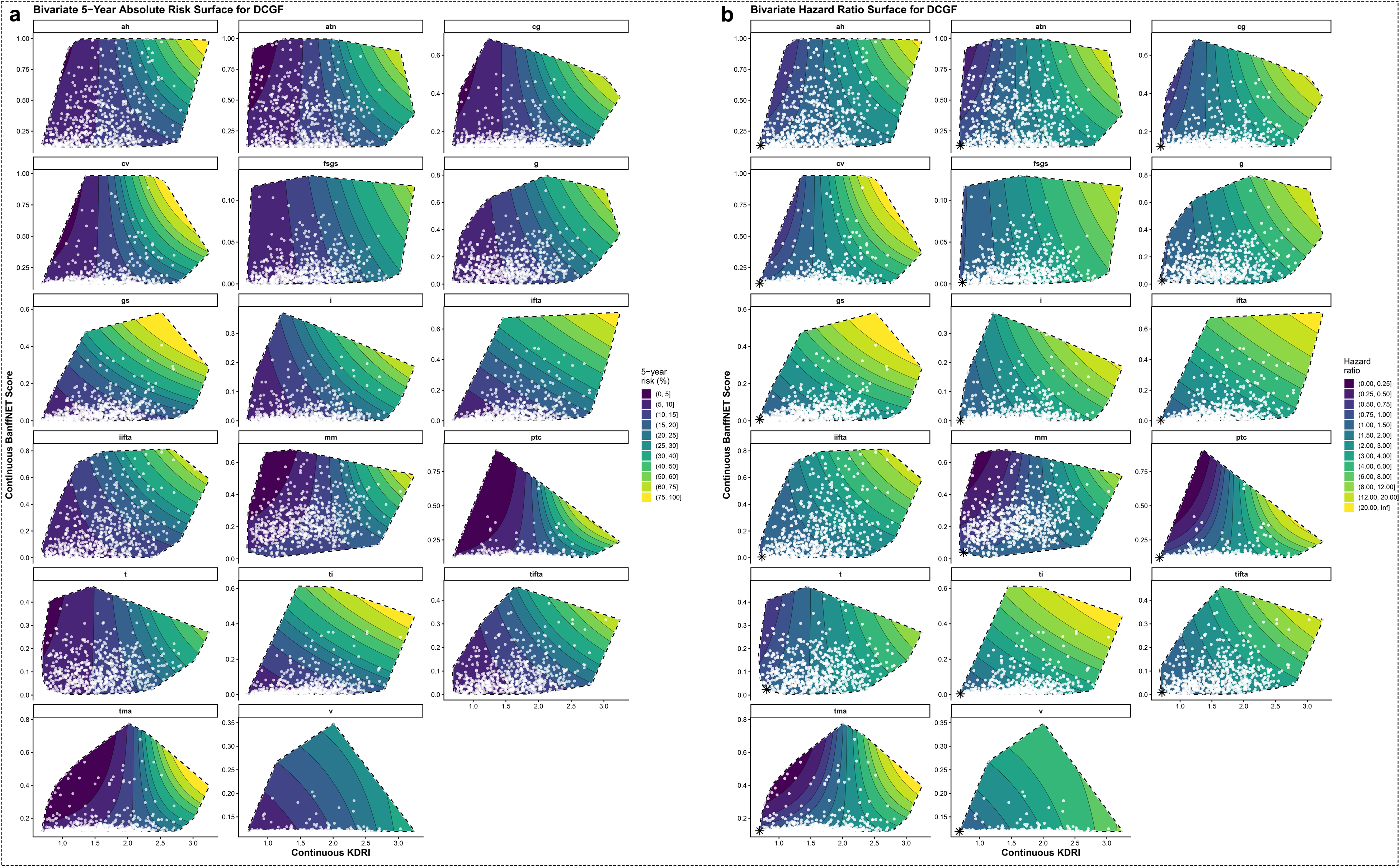
Bivariate associations of KDRI and individual BanffNET scores with death-censored graft failure (DCGF). Bivariate surface plots illustrate the joint associations of KDRI (x-axis) and BanffNET score (y-axis) with transplant outcomes. (a) Predicted 5-year absolute risk of DCGF. (b) Hazard ratio for DCGF, relative to KDRI = 0.68 and BanffNET score = 1.05. Colored surfaces represent the magnitude of the predicted probability, odds ratio, absolute risk, or hazard ratio according to the corresponding color scales, with contour lines indicating levels of equal predicted outcome or relative effect. White points show the observed distribution of KDRI and BanffNET score in the study population. Dashed boundaries delineate the region of the covariate space supported by the observed data.

## Notes

### Competing Interest Statement

The authors have declared no competing interest.

### Author Declarations

Leiden University Medical Center (LUMC) Review Committee Biobank & Biomaterials (TCBio). This study falls under the approval of the Non-WMO Committee of Leiden University Medical Center, Department of Internal Medicine (ID138026).

