## Supplementary Tables for "Validation of the Incremental Prognostic Value of Deceased Donation Pre-implantation Kidney Transplant Biopsies through Comprehensive Lesion Quantification with Deep Learning"

**Supplementary Table 1. Overview of variables included in the predefined covariate sets**

| Covariate sets | Variables |
|---|---|
| **Set 1** | Donor characteristic: KDRI<br>Recipient characteristics: age, sex, height, weight, history of diabetes and hypertension, prior kidney transplantation, HLA mismatch level, and the presence of DSA prior to transplantation |
| **Set 2** | **Set 1** incorporates Pathologist-assessed lesion scores: g, mm, i, t, ti, ifta, iifta, tifta, cv, ah, tma, and ggs |
| **Set 3** | **Set 1** incorporates BanffNET scores: g, cg, mm, i, t, ti, ifta, iifta, tifta, ptc, v, cv, ah, tma, ati, ggs and fsgs |
| **Set 4** | **Set 1** incorporates the first seven PCs derived from Pathologist-assessed lesion scores |
| **Set 5** | **Set 1** incorporates the first seven PCs derived from BanffNET scores |

KDRI, kidney donor risk index; DSA, donor-specific antibodies; g: glomerulitis; cg, transplant glomerulopathy; mm, mesangial matrix expansion; i, interstitial inflammation; t, tubulitis; ti, total inflammation; ifta, interstitial fibrosis and tubular atrophy; iifta, interstitial inflammation in areas of IFTA; tifta, tubulitis in areas of IFTA; ptc, peritubular capillaritis; v, intimal arteritis; cv, vascular fibrous Intimal thickening; ah, arteriolar hyalinosis; tma, thrombotic microangiopathy; and ati, acute tubular injury; ggs, global glomerulosclerosis, defined as the percentage of glomeruli sclerosed; fsgs, defined as the percentage of glomeruli affected by focal and segmental sclerosis.

**Supplementary Table 2. Summary of delta AUC relative to baseline across outcomes and biomarkers**

| Outcome / Biomarker | Model | Mean ± SD | Median [IQR] |
|---|---|---|---|
| Indication biopsy - eGFR | Baseline | | |
| | Baseline + BanffNET | 0.04 ± 0.00 | 0.04 [0.04–0.04] |
| | Baseline + BanffNET (PC) | 0.04 ± 0.01 | 0.04 [0.03–0.04] |
| | Baseline + Pathologist | 0.04 ± 0.01 | 0.04 [0.03–0.04] |
| | Baseline + Pathologist (PC) | 0.02 ± 0.01 | 0.02 [0.01–0.02] |
| Indication biopsy - UPCR | Baseline | | |
| | Baseline + BanffNET | 0.06 ± 0.01 | 0.06 [0.06–0.07] |
| | Baseline + BanffNET (PC) | 0.03 ± 0.01 | 0.04 [0.03–0.04] |
| | Baseline + Pathologist | 0.03 ± 0.01 | 0.03 [0.03–0.04] |
| | Baseline + Pathologist (PC) | 0.02 ± 0.01 | 0.02 [0.01–0.02] |
| DCGF - eGFR | Baseline | | |
| | Baseline + BanffNET | -0.00 ± 0.01 | -0.00 [-0.01–0.00] |
| | Baseline + BanffNET (PC) | 0.00 ± 0.01 | -0.00 [-0.00–0.01] |
| | Baseline + Pathologist | 0.03 ± 0.01 | 0.02 [0.02–0.03] |
| | Baseline + Pathologist (PC) | 0.01 ± 0.01 | 0.01 [0.01–0.02] |
| DCGF - UPCR | Baseline | | |
| | Baseline + BanffNET | 0.00 ± 0.01 | 0.00 [-0.01–0.01] |
| | Baseline + BanffNET (PC) | 0.01 ± 0.01 | 0.00 [0.00–0.01] |
| | Baseline + Pathologist | 0.04 ± 0.01 | 0.03 [0.03–0.04] |
| | Baseline + Pathologist (PC) | 0.02 ± 0.01 | 0.02 [0.01–0.02] |

Mean **Δ**AUC was calculated as the average yearly change in AUC over a 10-year prediction horizon

**Supplementary Table 3. Summary of delta Brier relative to baseline across outcomes and biomarkers**

| Outcome / Biomarker | Model | Mean ± SD | Median [IQR] |
|---|---|---|---|
| Indication biopsy - eGFR | Baseline | | |
| | Baseline + BanffNET | -0.02 ± 0.00 | -0.02 [-0.02–-0.02] |
| | Baseline + BanffNET (PC) | -0.02 ± 0.00 | -0.02 [-0.02–-0.02] |
| | Baseline + Pathologist | -0.04 ± 0.01 | -0.04 [-0.04–-0.03] |
| | Baseline + Pathologist (PC) | -0.01 ± 0.00 | -0.01 [-0.02–-0.01] |
| Indication biopsy - UPCR | Baseline | | |
| | Baseline + BanffNET | -0.01 ± 0.00 | -0.01 [-0.01–-0.01] |
| | Baseline + BanffNET (PC) | -0.00 ± 0.00 | -0.00 [-0.00–-0.00] |
| | Baseline + Pathologist | -0.01 ± 0.00 | -0.01 [-0.01–-0.01] |
| | Baseline + Pathologist (PC) | -0.00 ± 0.00 | -0.00 [-0.00–-0.00] |
| DCGF - eGFR | Baseline | | |
| | Baseline + BanffNET | -0.02 ± 0.01 | -0.02 [-0.03–-0.02] |
| | Baseline + BanffNET (PC) | -0.01 ± 0.00 | -0.01 [-0.01–-0.01] |
| | Baseline + Pathologist | -0.01 ± 0.00 | -0.01 [-0.01–-0.00] |
| | Baseline + Pathologist (PC) | -0.01 ± 0.00 | -0.01 [-0.01–-0.00] |
| DCGF - UPCR | Baseline | | |
| | Baseline + BanffNET | 0.00 ± 0.00 | 0.00 [-0.00–0.01] |
| | Baseline + BanffNET (PC) | -0.00 ± 0.00 | -0.00 [-0.00–-0.00] |
| | Baseline + Pathologist | -0.00 ± 0.00 | -0.00 [-0.01–-0.00] |
| | Baseline + Pathologist (PC) | -0.00 ± 0.00 | -0.00 [-0.00–-0.00] |

Mean **Δ**Brier was calculated as the average yearly change in Brier over a 10-year prediction horizon
